# The Effectiveness and Cost-Effectiveness of CashPlus Interventions to Prevent Acute Malnutrition in Somalia: Evidence from an Adaptive Cluster Randomized Control Trial

**DOI:** 10.1101/2025.07.22.25331867

**Authors:** Nadia Akseer, Kemish Kenneth Alier, Samantha Grounds, Sydney Garretson, Sagal Mohamud, Qundeel Khattak, Marina Tripaldi, Fabrizio Loddo, Said Aden Mohamoud, Adan Yusuf Mahdi, Mohamoud Ali Nur, Sadiq Mohamed Abdiqadir, Emily Mitchell, Andreas Kees, Mohamed Billow Mahat, Maimun Gure, Dahir Isaq Jibril, Dahir Gedi, Michael Ocircan P’Rajom, Meftuh Omer, Abdullahi Farah, Mohamed Abdirashid Osman, Abdiaziz Mohamed Adan, Farhan Mohamed Mohamoud, Abdullahi Muse Mohamoud, Abdifatah Ahmed Mohamed, Abdulkadir Ali Abdi, Adam Abdulkadir, Shelley Walton

## Abstract

**Background:** Acute malnutrition affects millions of children under five (U5) and pregnant and lactating women (PLW) globally, especially in humanitarian settings. Although “CashPlus” interventions – cash or food transfers combined with complementary components – are widely implemented, evidence remains limited on which combinations and durations are most effective at preventing acute malnutrition.

**Methods:** We conducted a three-arm cluster-randomized trial within Save the Children’s ‘CashPlus for Nutrition’ program in Somalia. Monthly support was provided as: (1) cash alone (Arm 1), (2) cash plus social and behavior change communication (SBCC; Arm 2), or (3) cash plus an additional cash top-up (Arm 3). Thirty-three villages were randomized across arms, targeting ∼1,500 households. Primary outcomes included prevalence and incidence of acute malnutrition in children 6–59 months and mothers, assessed at baseline, midline (3 months), and endline (6 months), alongside economic evaluations of cost and cost-effectiveness. Market monitoring and qualitative data were also conducted.

**Results:** Child acute malnutrition prevalence was approximately 15% in each arm at baseline. After 3 months, prevalence declined by 2.0 percentage points in Arm 1 to 13.0% (95% CI: 10.3%–16.1%), representing a relative reduction of 13.3%. In Arm 2, prevalence declined by 5.9 percentage points to 9.1% (6.8%–11.8%), a relative reduction of 39.3%. In contrast, prevalence in Arm 3 remained essentially unchanged, with an increase to 15.1% (12.1%–18.6%). By endline, there was little change from midline in all arms.

Maternal malnutrition improved most in Arm 2, but differences were not statistically significant. All arms showed improvements in dietary diversity and food security, but only Arm 2 achieved sustained nutrition gains. Household livelihood conditions appeared to improve overall though monthly expenditures nearly doubled. Arm 2 was the most effective and cost effective.

**Conclusion:** Adding SBCC to cash transfers significantly improved child nutrition compared to cash alone, highlighting that integrated approaches can enhance nutritional outcomes and cost-effectiveness in humanitarian settings.

**Registration:** The cluster-RCT was registered retrospectively at ClinicalTrials.gov on October 14, 2025, ID: NCT06642012.

## Introduction

Somalia, an impoverished country that has experienced years of conflict, violence and environmental disasters, continues to suffer from complex emergencies that result in large-scale population displacement, disease and food insecurity.[1] Recent humanitarian aid in Somalia has focused on relief from extreme drought between 2020 and 2022, and severe flooding in 2023, which has reduced agricultural production and damaged critical healthcare and water, sanitation, and hygiene (WaSH) infrastructure.[1] Despite the ongoing humanitarian presence, the persisting conflict and climate disasters contribute to high rates of food insecurity throughout Somalia.[2] The Integrated Food Security Phase Classification (IPC) estimated that 4 million people (21% of Somalia’s population) were living in Crisis or Emergency states of food insecurity from January to March 2024.[3]

Children under five years old (CU5) are especially vulnerable to the impacts of food insecurity due to increased nutritional needs during this critical period of child development; and food insecure children have heightened risk of acute malnutrition (also referred to as wasting throughout this paper), morbidity and mortality. [4] The latest estimates suggest that wasting prevalence among CU5 in Somalia ranges between 11.0% to 14.3% – more than double the global wasting rate of 6.8%, although these figures need updating in light of recent events.[5][6] The IPC projects that 1.7 million CU5 (approximately 45% of all CU5) in Somalia will suffer from acute malnutrition from January to December 2024.[3]

In 2023, USAID’s Bureau for Humanitarian Assistance (USAID/BHA) provided relief partners in Somalia with $761 million USD in funding for food and cash assistance, medical services and wasting treatment, and WaSH support.[2] In the United Nations Office for the Coordination of Humanitarian Affairs (OCHA)’s 2024 Humanitarian Needs and Response Plan, multi-purpose cash (MPC) programming was established as the desired modality for flexibly meeting Somali households’ basic needs.[7] In 2023, an estimated 8.3 million people in Somalia required this type of humanitarian assistance. [1,8]

Cash and Vouchers Assistance (CVA), also known as cash transfers, is increasingly used to address household food security and nutrition in humanitarian settings such as Yemen, Niger, and Somalia.[4,9–19] However, the evidence on the effectiveness and cost-effectiveness of various combinations and components of these interventions for prevention of wasting, especially when being implemented in such complex and fragile settings, needs further study.[9,11,12] CVA has proven to be cost-effective for improving food security, especially as mobile and direct delivery payment mechanisms can help reducing logistical costs, making cash a viable option even in challenging humanitarian contexts.[20,21] When combined with education or behavior change support, cash transfers can further enhance food security and dietary intake outcomes by enabling recipients to make informed choices about their food needs.[22] However, this existing evidence it‘s largely positive on intermediate outcomes such as food security and dietary diversity of individualsbut it’s mixed when it comes to nutrition outcomes such as prevention of acute malnutrition in humanitarian settings. [4,17,18,23] Additionally, as the role and importance of social and behavior change communication (SBCC) in complementing CVA has increasingly been recognized, there is also a need for further evidence around the effectiveness and cost-effectiveness of ‘cash plus SBCC’ approaches-such as the Resourcing families for Better Nutrition (in comparison to cash only interventions) for improving nutrition outcomes in humanitarian settings.[19,24–27]

Given these knowledge gaps, the need for additional research using randomized controlled trial design and mixed methods studies, to assess the impact and cost-effectiveness of various cash and ‘cash plus SBCC’ interventions in humanitarian settings has been emphasized.[4,12,13,15–19,23,28,29]

A few non-randomized trials conducted in Somalia measured the impact of CVA on CU5 nutrition outcomes[30,31], however these trials were limited in that they did not include a ‘cash plus’ SBCC intervention arm, explore the impact of different cash amounts, perform cost effectiveness analyses, or use WHZ as the gold standard for assessing child wasting. [32–35] The use of MUAC for assessing acute malnutrition may underestimate true wasting burden[31–34] More importantly, the trials lacked the rigor afforded by a randomized controlled trial design.[4,13] While a quasi-experimental trial in Somalia measured the impact of a ‘cash plus’ nutrition counseling intervention on child nutrition outcomes, the study did not conduct any cost analyses and lacked a ‘cash only’ intervention arm, preventing measuring the additional impact of SBCC when combined with cash in comparison to cash alone.[14] Moreover, market accessibility, functioning, and diversity are essential components of a successful cash for nutrition program because they ensure that households can purchase the necessary foods to meet their nutritional needs. Without functioning and diverse markets, cash transfers may not result in improved nutrition, as beneficiaries may lack access to nutritious food options or face inflated prices due to market inefficiencies. Previous trials have lacked the linkage of study impact and effects to availability and functionality of markets.

This study aimed to address the evidence gap on the effectiveness and cost-effectiveness of ‘cash plus’ approaches by undertaking a three-arm cluster-randomized trial embedded within a large ‘CashPlus for Nutrition’ program implemented by Save the Children in Somalia. The program is implemented in camps supporting internally displaced persons in two high-wasting burden regions of Somalia (Bay, Hiran). Specifically, the trial aims were:

1) To estimate and compare acute malnutrition incidence and prevalence of children 6-59 months-old and their mothers receiving on a monthly basis either cash (Arm 1 – control), cash + SBCC (Arm 2), or cash+ top-up cash (Arm 3); after 3 months and 6 months of cash assistance
2) To calculate the costs and cost-effectiveness of the different intervention arms
3) To understand determinants that may influence the effectiveness of the program: perspectives and experiences of mothers and fathers of CU5 beneficiaries of the cash program and the functionality of markets and food prices

The ongoing food insecurity crisis in Somalia necessitates continued humanitarian support and investigating cost-effective cash assistance interventions is of the utmost importance to implementers and program partners. Enhancing this evidence base will promote evidence-based decision-making and inform streamlined programmatic approaches for addressing malnutrition in Somalia and other humanitarian contexts.

## Methods

### Study Design

This study used a mixed method design inclusive of a prospective, adaptive, cluster-randomized controlled trial, qualitative data collection, market monitoring and costing analyses including cost-efficiency and cost-effectiveness.[36] An adaptive design was adopted for the trial to enable flexibility to adjust design and other parameters as needed in the complex, unpredictable humanitarian setting.[37–39] The UNICEF Conceptual Framework on Maternal and Child Nutrition categorizes the determinants of malnutrition into three groups: immediate, underlying, and basic/enabling causes.[27,28] This evidence-based conceptual framework informed the trial design and both quantitative and qualitative data collection and analysis.

### Study Context

Found in the Horn of Africa, Somalia experiences an arid/semi-arid climate defined by four alternating rainy and dry seasons. [42,43] This study took place in Baidoa (capital of the Bay region) and three districts of the Hiran region in central Somalia: Beledweyne (regional capital), Mataban, and Mahas.Both Bay and Hiran are comprised of urban, rural, nomadic, and IDP populations, with the majority of the population in both settings being rural or nomadic. [44,45] Around a quarter of Somalia’s IDPs are hosted in Baidoa, with IDP populations experiencing the highest burden of acute malnutrition. [46,47]

Conflict, insecurity, droughts, and floods have affected Bay and Hiran for years, impacting livelihoods, affecting food security, contributing to displacement, and resulting in the high rates of acute malnutrition reported in these regions. [47–51] A timeline of relevant shocks affecting the study areas over the study time period is available in **Figure S1**.

### BHA CashPlus for Nutrition Program

Save the Children implemented the USAID/BHA-funded CashPlus for Nutrition program in the Bay and Hiran regions of Somalia, which included unconditional cash transfers (UCTs) and social behavior change communication interventions to families largely based in IDP settings. The Cash for Nutrition implementation model followed the Save the Children’s common approach “Resourcing Families for Better Nutrition (RF4BN)”, where unconditional cash is combined a comprehensive set of Social and Behavior SBCC interventions that are context specific and tailored to the needs of the target group. [27] The SBCC package designed for this research is summarized in **Appendix S1 and S2**. Further program details are available in our methods paper.[36]

### Trial Design

In consultation with key stakeholders and given evidence gaps, we implemented the following three arms, each using UCTs as the base intervention:

- **Arm 1:** UCT to targeted families – 1 mobile transfer per month for 6 consecutive months. Households (HHs) in Bay received $90 per month and HHs in Hiran received $70 per month.
- **Arm 2:** UCT with SBCC – Cash was same amount asArm 1, coupled with the SBCC package. The SBCC package was context specific and tailored to the group of pregnant and breastfeeding women and CU5. This package included interpersonal communication, such as one-to-one counseling for mothers of children under five, and bi-monthly group sessions of approximately 45–60 minutes through the Mother-to-Mother Support Groups (MtMSGs), which covered key topics related to nutrition, health, and community awareness raising campaigns. Participants were enrolled in MtMSGs for two cycles of three months each, totaling 12 sessions. Additionally, the implementing partner organized cooking demonstrations for the support groups, highlighting nutritious foods available in the community.Additionally, the implementing partner also organized cooking demonstrations for the support groups, highlighting nutritious foods that are available in the community. Further details provided in **Appendix S1 and S2** and the methodology paper [36].
- **Arm 3:** UCT with additional monthly nutrition-cash top up – 1 per month for 6 months. Households in Bay received $90 plus an additional $35 top-up ($125 total) per month and HHs in Hiran received $70 plus an additional $35 top-up ($105 total) per month.

The base cash transfer amounts were established by the National Cash Working Group based on the food Minimum Expenditure Basket (MEB). Cash amounts were calculated to meet 80% of a person’s food energy needs (kcals) per month and were based on the average number of household members. [34,49,50].The Arm 3 top-up cash transfer amount was calculated using was aligned to the Cash Top up provide by WFP for nutrition; it was calculated by WFP using the Fill the Nutrient gap methodology to cover the additional nutritional needs of PLW and CU5, considering the available nutritious food in the project targeted locations and households’ capacity to meet their food needs.

### Study Population, Randomization and Sample Size

The trial randomized clusters (distinct villages) to one of three arms and then selected HHs within those arms as follows. An initial 44 clusters were identified for randomization which was then reduced to 33 based on accessibility and feasibility for research. The clusters were randomized to one of three arms using a random number generator. HHs with CU5 were identified from the BHA program roster and were contacted to enroll in the study. Additional details available in the methods paper.[36]

We estimated the number of HHs needed for the trial using child wasting as the primary outcome, considering feasibility/logistics in study settings, and a 7% minimum detectable difference in post-intervention prevalence of wasting. We assumed: 1) 20% baseline wasting 2) intra-class correlation coefficient (ICC) from earlier studies (0.02) [31,54]; average cluster size (150 HHs); 4) number of clusters (33 total, 11 per arm); 5) 5% significance and 80% power; and 6) CU5 per household (1.3 children). These parameters produced 410 HHs required per intervention group or 533 children. Accounting for 15% attrition, the final required sample size was 471 HHs or 613 children per arm. A total of 4,838 total HHs were evaluated for eligibility against the trial’s inclusion and exclusion criteria. 3,348 HHs were excluded because they did not have CU5 in the household or did not meet one or more exclusion criteria (**Box 1**). The final sample included 1490 HHs with 1,894 children, approximately 569-672 children across the arms (**Figure 1**).

**Figure 1.**
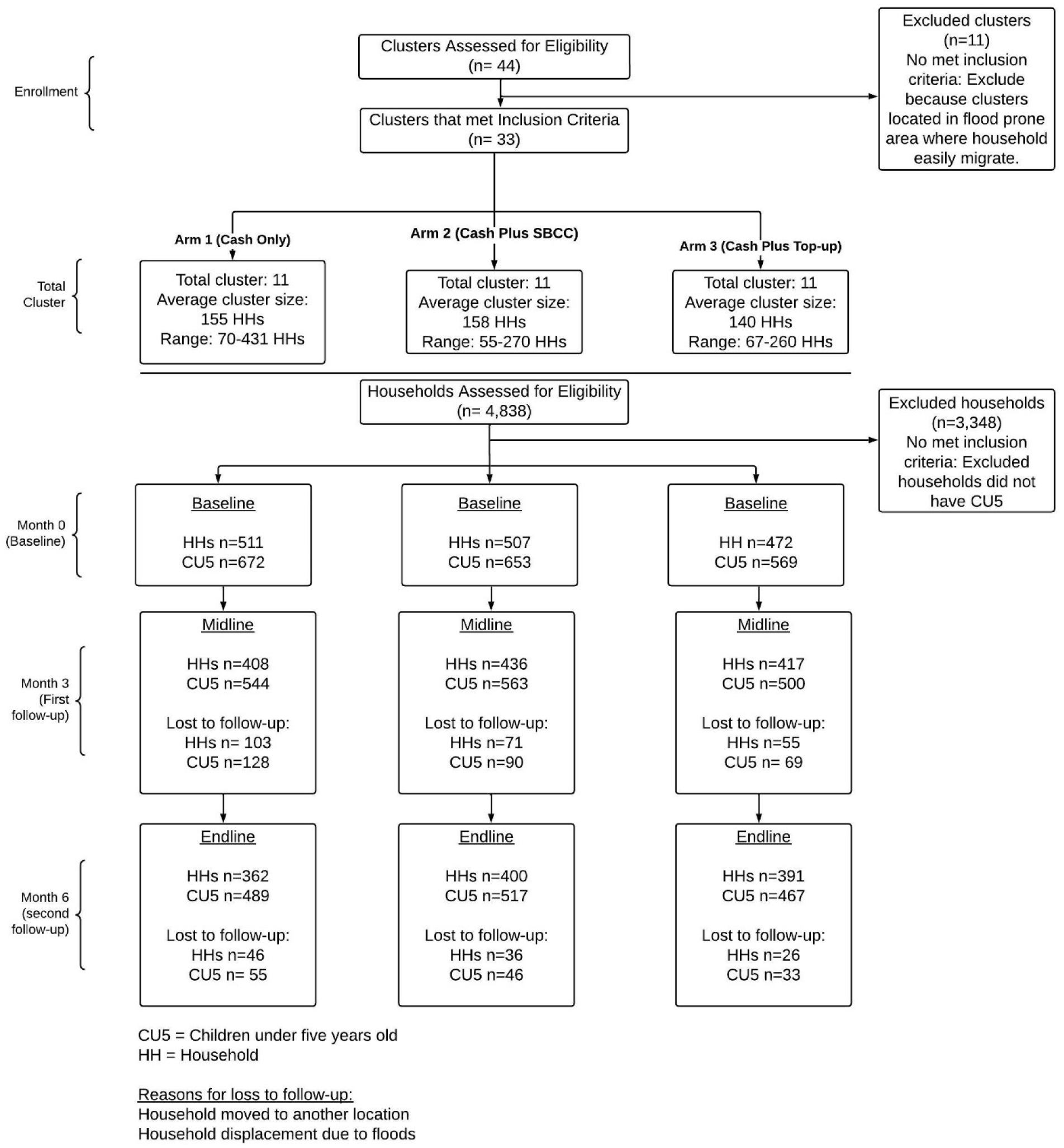
CONSORT Flowchart for Study Enrollment and Follow-Up.

**Box 1.**
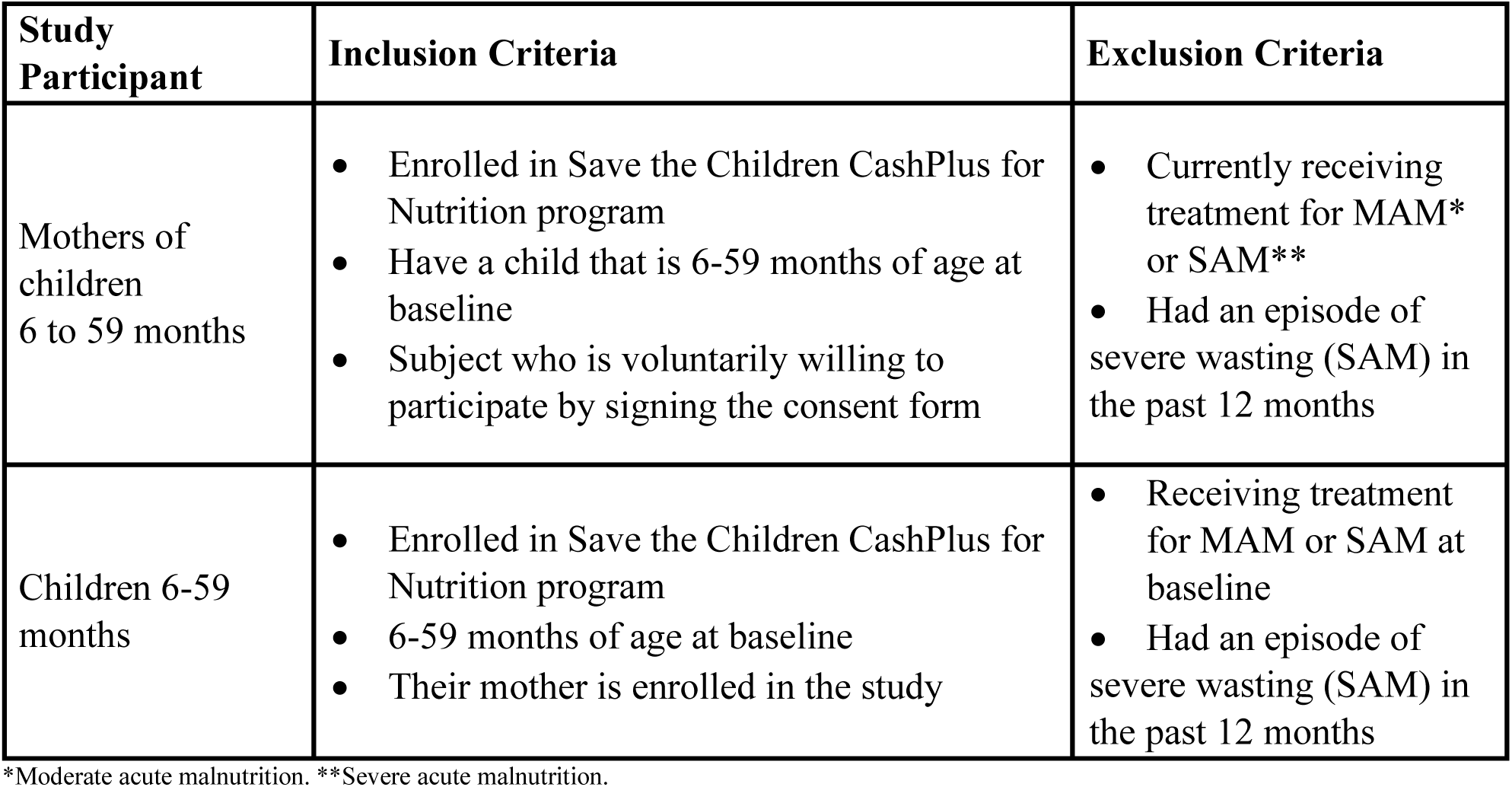
Trial Inclusion and Exclusion Criteria.

### Outcomes

The primary outcomes of interest were child wasting, defined by mid-upper arm circumference (MUAC), weight-for-height Z-score (WHZ), and bilaterial pitting edema; and maternal wasting, defined by MUAC. WHZ was calculated according to the 2006 WHO Child Growth Standards.[55–57] Values outside the WHO plausible ranges were coded as missing: WHZ < –5 or > 5, WAZ < –6 or > 5, and HAZ < –6 or > 6. Child wasting was categorized as a binary outcome (wasted/not wasted), and as a categorical outcome, with wasting stratified into moderate (MAM) or severe wasting (SAM). Maternal wasting was categorized as overweight, normal, or moderate acute malnutrition. [58] A secondary outcome of interest was child stunting, defined by height-for-age Z-score (HAZ). For children who exceeded 59 months of age during midline and endline follow-up, anthropometric measurements were standardized at 59 months. [52–54] Full outcome definitions can be found in **Appendix S3**.

### Data Collection and Analysis

The trial was implemented from June to November 2023. Baseline quantitative data collection occurred in May 2023, midline occurred in September 2023, and endline occurred in December 2023. Qualitative data were collected in January 2024. Local enumerators were recruited and underwent comprehensive training prior to baseline data collection. Standard procedures and equipment were used to collect data (details in **Appendix S4**).

Along with outcome data, the quantitative survey instrument implemented at each timepoint collected information on immediate, underlying, and enabling causes of malnutrition.[59,60] A comprehensive list of indicators and definitions can be found in the **Appendix S5**.

This study employed a mixed-methods approach encompassing quantitative, qualitative, economic, and market analyses, with further details provided in **Appendices S6-S11** and our methods paper [36]. Quantitative data were rigorously cleaned and validated in near real-time, with main analyses conducted using an intention-to-treat (ITT) approach and complemented by per-protocol (PP) and incidence analyses to test the robustness of findings. Multilevel mixed-effects models were used to estimate treatment effects, supported by a difference-in-differences logistic regression framework and sensitivity analyses addressing missing data, time specifications, and loss to follow-up. Adjusted predictions and marginal effects were calculated, and all models adhered to CONSORT guidelines (**Appendix S6)**. Qualitative data were collected through focus group discussions with mothers and fathers to contextualize quantitative findings, using a sequential explanatory design (**Appendix S6, S7**). Cost-efficiency and cost-effectiveness analyses triangulated findings from financial records, modeling, and staff consultations, applying a top-down, activity-based costing approach [61,62] that included societal costs (**Appendices F, H, I**). Market monitoring was conducted at three time points using vendor surveys in both regions to track the availability and prices of key food commodities, enabling analysis of how contextual shifts may have influenced household behavior and intervention impacts (**Appendices S6, S10, S11**).

## Results

We present the results first by describing the sample followed by the key maternal and child outcomes (effectiveness and cost-effectiveness) and then delve into exploring potential factors that may influence the outcomes including perspectives from beneficiaries and market functionality.

### Baseline Descriptive

Baseline descriptive statistics of the study population are presented in **Table 1**. The sample comprised one-third from Bay and two-thirds from Hiran, with a similar distribution between IDP settings (one-third) and host communities (two-thirds). Arm 3 included slightly more participants from Bay and a higher proportion of IDPs.

**Table 1.** Baseline Characteristics of Households, Mother and Children.

| Characteristics | Trial Arm |  |  |  |
| --- | --- | --- | --- | --- |
|  | Arm 1 (Cash Only)<br>(n=672) | Arm 2 (Cash Plus<br>SBCC)<br>(n=653) | Arm 3 (Cash Plus<br>Cash<br>top-up) (n=569) | p-value |
| Region |  |  |  |  |
| Bay | 252 (37.5%) | 250 (38.3%) | 295 (51.8%) | <0.001 |
| Hiran | 420 (62.5%) | 403 (61.7%) | 274 (48.2%) |  |
| Displacement status |  |  |  |  |
| Host | 416 (61.9%) | 403 (61.7%) | 271 (47.7%) | <0.001 |
| IDP | 256 (38.1%) | 250 (38.3%) | 297 (52.3%) |  |
| **Child Characteristics** |  |  |  |  |
| Child Sex |  |  |  |  |
| Male | 357 (53.1%) | 318 (48.7%) | 288 (50.6%) | 0.271 |
| Female | 315 (46.9%) | 335 (51.3%) | 281 (49.4%) |  |
| Age, months | 34.4 (14.6) | 34.7 (14.3) | 33.7 (13.9) | 0.507 |
| Age (Dichotomized) |  |  |  |  |
| <2 years | 162 (24.1%) | 150 (23.0%) | 134 (23.6%) | 0.888 |
| 2+ years | 510 (75.9%) | 503 (77.0%) | 435 (76.4%) |  |
| Child MUAC | 14.3 (1.0) | 14.5 (1.0) | 14.3 (1.2) | 0.069 |
| WHZ-Score | -0.9 (1.1) | -0.9 (1.0) | -0.9 (1.1) | 0.963 |
| WAZ-Score | -1.2 (0.9) | -1.1 (0.9) | -1.3 (0.9) | 0.047 |
| HAZ-Score | -1.0 (1.3) | -1.0 (1.3) | -1.2 (1.4) | 0.007 |
| Wasting (WHZ) |  |  |  |  |
| WHZ ≥ -2 SD | 567 (85.3%) | 554 (85.0%) | 480 (84.5%) | 0.933 |
| WHZ < -2 SD | 98 (14.7%) | 98 (15.0%) | 88 (15.5%) |  |
| Wasting (MUAC) |  |  |  |  |
| MUAC ≥ 11.5 cm | 670 (99.7%) | 652 (100.0%) | 567 (99.6%) | 0.342 |
| MUAC < 11.5 cm | 2 (0.3%) | 0 (0.0%) | 2 (0.4%) |  |
| Underweight (WAZ) |  |  |  |  |
| WAZ ≥ -2 SD | 554 (82.7%) | 556 (85.1%) | 455 (80.0%) | 0.058 |
| WAZ < -2 SD | 116 (17.3%) | 97 (14.9%) | 114 (20.0%) |  |
| Stunting (HAZ) |  |  |  |  |
| HAZ ≥ -2 SD | 537 (80.4%) | 534 (82.0%) | 410 (72.3%) | <0.001 |
| HAZ <-2 SD | 131 (19.6%) | 117 (18.0%) | 157 (27.7%) |  |
| Child ever breastfed<br>(youngest child only 6-23mo) |  |  |  |  |
| No | 18 (10.3%) | 27 (16.9%) | 18 (12.6%) | 0.198 |
| Yes | 157 (89.7%) | 133 (83.12%) | 125 (87.4%) |  |
| Received any childhood<br>vaccination+ |  |  |  |  |
| No | 86 (49.1%) | 82 (51.6%) | 77 (53.8%) | 0.704 |
| Yes | 89 (50.9%) | 77 (48.4%) | 66 (46.2%) |  |
| Had childhood illness 2<br>weeks prior |  |  |  |  |
| No | 103 (58.9%) | 109 (68.6%) | 106 (74.1%) | 0.013 |
| Yes | 72 (41.1%) | 50 (31.4%) | 37 (25.9%) |  |
| Child Minimum Dietary<br>Diversity (MDD-C)* |  |  |  |  |
| Do Not Meet MDD | 149 (94.9%) | 117 (88.0%) | 112 (89.6%) | 0.093 |
| Meet MDD | 8 (5.1%) | 16 (12.0%) | 13 (10.4%) |  |
| Animal-sourced protein<br>(meat, eggs, milk) |  |  |  |  |
| No animal-sourced<br>protein | 82 (46.9%) | 66 (41.3%) | 62 (43.4%) | 0.579 |
| At least one animal-sourced protein | 93 (53.1%) | 94 (58.7%) | 81 (56.6%) |  |
| **Mother Characteristics**<br>**(n=1490)** |  |  |  |  |
| Age, years | 32.2 (9.0) | 34 (10.8) | 32.9 (9.9) | 0.014 |
| Age (Dichotomized) |  |  |  |  |
| 18-34 Yrs | 306 (59.9%) | 281 (55.5%) | 278 (59.2%) | 0.325 |
| 35+ Yrs | 205 (40.1%) | 225 (44.5%) | 192 (40.8%) |  |
| MUAC, cm | 26.7 (4.0) | 27.0 (3.8) | 26.1 (3.6) | 0.002 |
| Maternal Wasting (MUAC) |  |  |  |  |
| MUAC < 23 cm | 67 (13.9%) | 55 (11.5%) | 69 (15.5%) | 0.193 |
| MUAC ≥ 23 cm | 415 (86.1%) | 424 (88.5%) | 375 (84.5%) |  |
| Pregnancy status |  |  |  |  |
| No | 420 (82.2%) | 400 (79.1%) | 370 (78.4%) | 0.277 |
| Yes | 91 (17.8%) | 106 (20.9%) | 102 (21.6%) |  |
| Education level |  |  |  |  |
| No Formal Education | 420 (82.2%) | 418 (82.6%) | 410 (86.9%) | 0.215 |
| Madrasa | 51 (10.0%) | 51 (10.1%) | 40 (8.5%) |  |
| Primary & Secondary | 40 (7.8%) | 37 (7.3%) | 22 (4.7%) |  |

| Household Characteristics (n=1490) |  |  |  |  |
| --- | --- | --- | --- | --- |
| Mother head of household |  |  |  |  |
| No | 360 (70.5%) | 354 (70.0%) | 339 (71.8%) | 0.804 |
| Yes | 151 (29.5%) | 152 (30.0%) | 133 (28.2%) |  |
| Decision-making on income |  |  |  |  |
| Jointly | 215 (42.5%) | 205 (41.0%) | 199 (42.4%) | 0.95 |
| Mother | 111 (21.9%) | 117 (23.4%) | 100 (21.3%) |  |
| Father | 180 (35.6%) | 178 (35.6%) | 170 (36.3%) |  |
| Decision-making on healthcare |  |  |  |  |
| Jointly | 248 (48.9%) | 266 (53.1%) | 254 (54.2%) | 0.188 |
| Mother | 100 (19.7%) | 109 (21.8%) | 90 (19.2%) |  |
| Father | 159 (31.4%) | 126 (25.2%) | 125 (26.7%) |  |
| Number of U5 children in the household |  |  |  |  |
| 1 child | 351 (68.7%) | 367 (72.4%) | 373 (79.0%) | 0.001 |
| ≥ 2 children | 160 (31.3%) | 140 (27.6%) | 99 (21.0%) |  |
| Household Hunger Scale (0 best-6 worse) |  |  |  |  |
| Little to no Hunger (≤ 1 score) | 283 (55.4%) | 307 (60.7%) | 307 (65.0%) | 0.001 |
| Moderate Hunger (2-3 score) | 219 (42.9%) | 197 (38.9%) | 165 (35.0%) |  |
| Severe Hunger (4-6 score) | 9 (1.7%) | 2 (0.4%) | 0 (0.0%) |  |
| Food Consumption Score (adjusted for high oils & sugar consumption) |  |  |  |  |
| Poor (≤ 28 score) | 262 (51.3%) | 231 (45.6%) | 189 (40.0%) | 0.007 |
| Borderline (28.5-42 score) | 124 (24.3%) | 131 (25.9%) | 125 (26.5%) |  |
| Acceptable (≥ 43 score) | 125 (24.4%) | 144 (28.5%) | 158 (33.5%) |  |
| Reduced Coping Strategies Index Score (0 best-56 worse)** | 14.5 (9.0) | 15.3 (8.7) | 14.9 (9.4) | 0.368 |
| Monthly Expenditure, Total (USD) | 113.1 (55.5) | 111.6 (69.4) | 95 (53.3) | <0.001 |
| Percent of Expenditure on Food (%) | 65.7% | 70.6% | 68.8% | <0.001 |
| Crowding |  |  |  |  |
| Not Crowded (<5 household members) | 165 (32.3%) | 175 (34.5%) | 212 (44.9%) | <0.001 |
| Crowded ($\geq 5$ household members) | 346 (67.7%) | 332 (65.5%) | 260 (55.1%) | |
| Sanitation |  |  |  |  |
| Open defecation | 120 (23.5%) | 69 (13.7%) | 100 (21.3%) | <0.001 |
| Flush/Latrine | 390 (76.5%) | 436 (86.3%) | 370 (78.7%) |  |
| Drinking water source+ |  |  |  |  |
| Unprotected Source | 209 (41.4%) | 163 (32.3%) | 205 (43.5%) | 0.001 |
| Protected source | 299 (58.9%) | 341 (67.7%) | 266 (56.5%) |  |

1) Response rate for the different variables varied
2) Questions on breastfeeding, vaccination, childhood illnesses and dietary diversity were only asked to the youngest child in the household
\* Minimum Dietary Diversity for the Child
\*\* Reduced Coping strategies Index (rCSI): Higher scores mean frequent use of harmful coping strategies and hence increased food insecurity.
+ Protected water source: piped/tap water and borehole; Unprotected water source: well, rainwater, tanker truck, rain, and surface water
n=number of U5 children

Overall, study arms were balanced in key maternal and child characteristics. Child wasting was approximately 15% at baseline, with no statistically significant differences across arms. Children in Arm 3 had slightly higher stunting rates but fewer recent illnesses. Mothers’ average age (32– 34 years) and MUAC (26–27 cm) showed minor variation across arms.

Household crowding was lower in Arm 3, with fewer children and total household members, and food security indicators were better compared to other arms. Average monthly expenditures were lower in Arm 3 (USD 95) than in other arms (USD 112–113), with all households spending about 70% on food.

Most households had access to flush or latrine toilets (77%–86%, highest in Arm 2), and 57%– 68% had access to protected water sources, also highest in Arm 2. Similar midline and endline analyses are available in **Table S1**.

Across the 6-month follow-up period, the study experienced 22% attrition overall; this varied slightly across arms (18%-27%) (**Figure 1**). However, children that were retained compared to those that were lost to follow-up were similar in demographics and nutritional outcomes at both midline and endline (**Table S2**).

### Maternal and Child Outcomes

Using ITT analyses, from baseline to midline, child wasting prevalence decreased significantly in Arm 2 from 15.0% [95%CI: 12.4%-18.0%] to 9.1% [95%CI: 6.8%-11.8%] but not in Arm 1 (14.7% to 13.0%; confidence intervals overlap) or Arm 3 (15.5% to 15.1%; confidence intervals overlap) (**Figure 2A**). Arm 2 wasting prevalence was statistically different from Arm 1 (p=0.039) and Arm 3 (p=0.003) at midline. By endline, all arms experienced a slight increase in wasting prevalence (15.0%, 10.1% and 17.4% for Arms 1-3, respectively) from midline levels but Arm 2 was statistically lower compared to other arms (p<0.05) and sustained the greatest reduction overall from baseline to endline. Analysis of wasting incidence outcomes found similar results (**Figure 2B**). Over-time and between-arm differences for child anthropometry (MUAC, WHZ, WAZ and HAZ) outcomes overall and by sub-group (Bay vs Hiran regions; child <2 vs child > = 2) are found in **Table S3** (prevalence) and **Table S4** (incidence).

**Figure 2.**
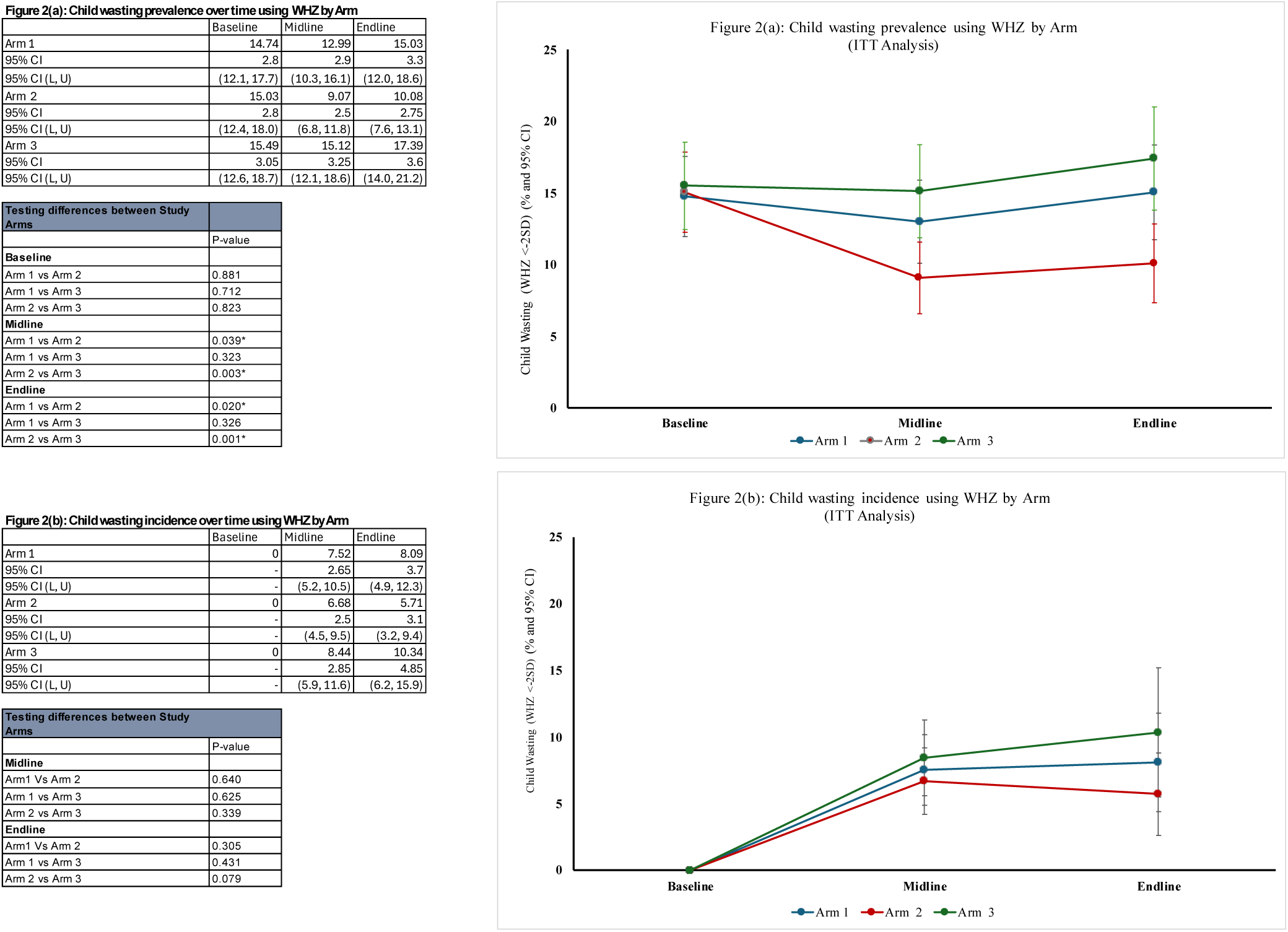
Child wasting prevalence and incidence over time using WHZ by Arm.

Sensitivity analyses comparing ITT and per-protocol findings showed no differences in child and maternal wasting prevalence trends but a slight underestimation of wasting incidence using PP methods (**Figure S2A, S2B**). Given these minor differences, we report wasting prevalence using ITT analysis for the remaining results.

Child wasting prevalence differences existed and even amplified after adjusting for baseline covariates to account for potential confounding (**Table 2**). Similar findings were seen for child underweight prevalence differences in Arm 2 compared to other arms, while no effect on stunting prevalence change was observed (**Table S5**). Crude and adjusted difference-in-difference analyses confirmed these findings, with protective odds ratios for child wasting in Arm 2 vs. Arm 1, but no difference between Arms 3 and 1 (**Table 3**). Difference-in-difference models for child underweight, stunting, and MUAC-based wasting showed no meaningful differences between arms in either crude or adjusted models (**Table 3**); similar models stratified by region and age group are shown in **Table S6**.

**Table 2.** Crude and adjusted differences for child wasting outcomes (Prevalence analysis using ITT Approach)

|  |  | Unadjusted |  |  |  |  |  | Partially Adjusted* |  |  |  |  |  | Fully Adjusted** |  |  |  |  |  |
| --- | --- | --- | --- | --- | --- | --- | --- | --- | --- | --- | --- | --- | --- | --- | --- | --- | --- | --- | --- |
| Outcome variable | Study Arm | Endline - Baseline | p-val | Midline - Baseline | p-val | Endline - Midline | p-val | Endline - Baseline | p-val | Midline - Baseline | p-val | Endline - Midline | p-val | Endline - Baseline | p-val | Midline - Baseline | p-val | Endline - Midline | p-val |
| Child Outcomes |  |  |  |  |  |  |  |  |  |  |  |  |  |  |  |  |  |  |  |
| WHZ (mean/95% CI) | Arm 1 | 0.111 (0.013, 0.210) | 0.026 | 0.165 (0.072, 0.259) | 0.001 | 0.074 (0.012, 0.137) | 0.020 | 0.263 (0.055, 0.472) | 0.013 | 0.449 (0.253, 0.645) | <0.001 | 0.185 (0.050, 0.321) | 0.007 | -0.048 (-0.310, 0.215) | 0.721 | 0.257 (0.006, 0.507) | 0.044 | 0.033 (-0.142, 0.208) | 0.712 |
|  | Arm 2 | 0.148 (0.052, 0.243) | 0.003 | 0.242 (0.151, 0.333) | <0.001 | 0.085 (0.023, 0.147) | 0.007 | 0.372 (0.095, 0.650) | 0.008 | 0.569 (0.309, 0.829) | <0.001 | 0.227 (0.047, 0.407) | 0.013 | 0.148 (-0.179, 0.474) | 0.375 | 0.467 (0.157, 0.777) | 0.003 | 0.116 (-0.102, 0.334) | 0.298 |
|  | Arm 3 | -0.010 (-0.110, 0.089) | 0.840 | 0.092 (-0.004, 0.187) | 0.059 | 0.011 (-0.054, 0.075) | 0.749 | 0.318 (-0.089, 0.725) | 0.126 | 0.477 (0.094, 0.860) | 0.015 | 0.200 (-0.068, 0.467) | 0.143 | 0.231 (-0.212, 0.674) | 0.307 | 0.519 (0.098, 0.940) | 0.016 | 0.154 (-0.144, 0.452) | 0.310 |
| MUAC, cm (mean/95% CI) | Arm 1 | 0.443 (0.352, 0.534) | <0.001 | 0.366 (0.280, 0.452) | <0.001 | 0.221 (0.166, 0.275) | <0.001 | 0.641 (0.435, 0.847) | <0.001 | 0.483 (0.291, 0.676) | <0.001 | 0.336 (0.216, 0.456) | <0.001 | 0.517 (0.254, 0.780) | <0.001 | 0.403 (0.155, 0.652) | 0.001 | 0.271 (0.114, 0.427) | 0.001 |
|  | Arm 2 | 0.380 (0.291, 0.470) | <0.001 | 0.303 (0.219, 0.387) | <0.001 | 0.201 (0.147, 0.255) | <0.001 | 0.729 (0.455, 1.003) | <0.001 | 0.515 (0.260, 0.770) | <0.001 | 0.408 (0.248, 0.568) | <0.001 | 0.578 (0.250, 0.905) | 0.001 | 0.396 (0.088, 0.704) | 0.012 | 0.327 (0.133, 0.522) | 0.001 |
|  | Arm 3 | 0.185 (0.093, 0.278) | <0.001 | 0.160 (0.072, 0.248) | <0.001 | 0.106 (0.049, 0.162) | <0.001 | 0.734 (0.332, 1.136) | <0.001 | 0.491 (0.115, 0.867) | 0.011 | 0.439 (0.201, 0.677) | <0.001 | 0.559 (0.114, 1.004) | 0.014 | 0.354 (-0.064, 0.772) | 0.097 | 0.343 (0.077, 0.609) | 0.011 |
| *Partially adjusted for cluster, child age and sex. |  |  |  |  |  |  |  |  |  |  |  |  |  |  |  |  |  |  |  |
| **Fully adjusted for cluster, child age, sex, displacement status, wasting , underweight, stunting at baseline, maternal education, HH monthly expenditure at baseline, sanitation and access to protected water source at baseline |  |  |  |  |  |  |  |  |  |  |  |  |  |  |  |  |  |  |  |

**Table 3.** Crude and adjusted logistic regression showing difference in differences effects for child anthropometry outcomes (Prevalence analysis using ITT Approach)

|  |  | Unadjusted model |  |  |  | Partially Adjusted model |  |  |  | Fully Adjusted model |  |  |  |
| --- | --- | --- | --- | --- | --- | --- | --- | --- | --- | --- | --- | --- | --- |
| Outcome variable | Study Arm | Midline | p-val | Endline | p-val | Midline | p-val | Endline | p-val | Midline | p-val | Endline | p-val |
| **Child Outcomes** |  | (n=1,607) |  | (n=1,473) |  | (n=1,607) |  | (n=1,473) |  | (n=1,607) |  | (n=1,473) |  |
| WHZ < -2 SD (OR, 95% CI) | Arm 2 | 0.57 (0.312, 1.034) | 0.064 | 0.53 (0.289, 0.972) | 0.040 | 0.53 (0.290, 0.980) | 0.043 | 0.52 (0.279, 0.956) | 0.035 | 0.55 (0.302, 1.017) | 0.057 | 0.52 (0.283, 0.967) | 0.039 |
|  | Arm 3 | 1.21 (0.671, 2.183) | 0.525 | 1.24 (0.687, 2.249) | 0.472 | 1.21 (0.667, 2.210) | 0.526 | 1.31 (0.720, 2.383) | 0.376 | 1.24 (0.677, 2.284) | 0.483 | 1.34 (0.730, 2.460) | 0.346 |
| WAZ < -2 SD (OR, 95% CI) | Arm 2 | 1.09 (0.550, 2.166) | 0.803 | 0.68 (0.334, 1.367) | 0.276 | 1.08 (0.543, 2.163) | 0.819 | 0.66 (0.325, 1.340) | 0.250 | 1.04 (0.510, 2.135) | 0.907 | 0.61 (0.293, 1.267) | 0.185 |
|  | Arm 3 | 1.35 (0.702, 2.585) | 0.370 | 1.62 (0.848, 3.099) | 0.144 | 1.36 (0.706, 2.620) | 0.359 | 1.62 (0.843, 3.107) | 0.148 | 1.24 (0.629, 2.447) | 0.535 | 1.54 (0.788, 3.021) | 0.206 |
| HAZ < -2 SD (OR, 95% CI) | Arm 2 | 1.23 (0.645, 2.330) | 0.534 | 0.83 (0.431, 1.600) | 0.578 | 1.17 (0.615, 2.226) | 0.632 | 0.78 (0.405, 1.507) | 0.461 | 1.14 (0.586, 2.199) | 0.707 | 0.75 (0.382, 1.468) | 0.400 |
|  | Arm 3 | 1.39 (0.754, 2.578) | 0.290 | 1.10 (0.588, 2.050) | 0.769 | 1.35 (0.730, 2.495) | 0.340 | 1.05 (0.564, 1.966) | 0.870 | 1.19 (0.629, 2.247) | 0.593 | 0.95 (0.499, 1.808) | 0.876 |
| MUAC < 11.5cm (OR, 95% CI) | Arm 2 | 1.59 (0.260, 9.730) | 0.616 | N/A | - | 1.63 (0.268, 9.912) | 0.595 | N/A | - | 1.72 (0.276, 10.740) | 0.561 | N/A | - |
|  | Arm 3 | 2.29 (0.210, 25.030) | 0.497 | 1.86 (0.184, 18.781) | 0.598 | 2.22 (0.205, 24.111) | 0.512 | 1.79 (0.178, 17.929) | 0.621 | 2.41 (0.214, 27.016) | 0.477 | 1.92 (0.184, 20.057) | 0.585 |

OR: Odd ratio; Arm 1 is the reference arm
N/A: No data point, omitted due to collinearity
n for child outcome is number of U5 children analyzed
Partially adjusted for cluster, child age, sex and wasting at baseline.
Fully adjusted for cluster, child age, sex, displacement status, wasting, underweight, stunting at baseline, maternal education HH monthly expenditure at baseline, sanitation and access to protected water source at baseline

Similar to the child, across all arms, maternal wasting prevalence decreased from baseline to midline but increased again at endline; Arm 2 had the lowest prevalence at endline (11.8% [95%CI:8.8%-15.3%]) compared to Arm 1 (15.5%) and Arm 3 (13.6%), though differences were not statistically significant (**Figure S3**). Over-time and between-arm differences for maternal MUAC overall and by sub-group are found in **Table S3** (prevalence) and **Table S4** (incidence). There were no differences across arms in maternal underweight after adjusting for covariates (**Table S5**).

### Cost Analysis

**Figure 3** and **Table S7** present the results of cost analyses for the program evaluation. Regarding the cost-efficiency of the intervention, Arms 1 and 2 provide the lowest cost per HH. Cash Transfer Ratio (CTR) was lower for Arm 3 given the nature of the intervention and larger cash transfer amount. When considering that Arm 2 received a comprehensive SBCC package, this arm appears to have little additional cost per HH over 6 months ($31) when compared to the other arms. In terms of overall reductions in wasting prevalence, compared to Arm 1, Arm 2 had 4.9% lower wasting prevalence by endline and Arm 3 had increased wasting by 1.9%. Overall, the CTRs for the three arms, which rang*e* from $0.69 to $0.80 of delivery costs per $1 in aid, are broadly in line with similar cash interventions in the region. The operational and administrative expenses of the SCI Somalia Country Office and SCI Global HQ – particularly staff salaries – were the main drivers of the CTR across all treatment arms. Direct costs such as logistics or cash transfer fees were not major cost drivers. Comparing Hiran and Bay, the project had lower delivery costs in Bay where the transfer value was $90, than in Hiran, where the transfer value was set at only $70 due to lower cost of living in the region.

**Figure 3.**
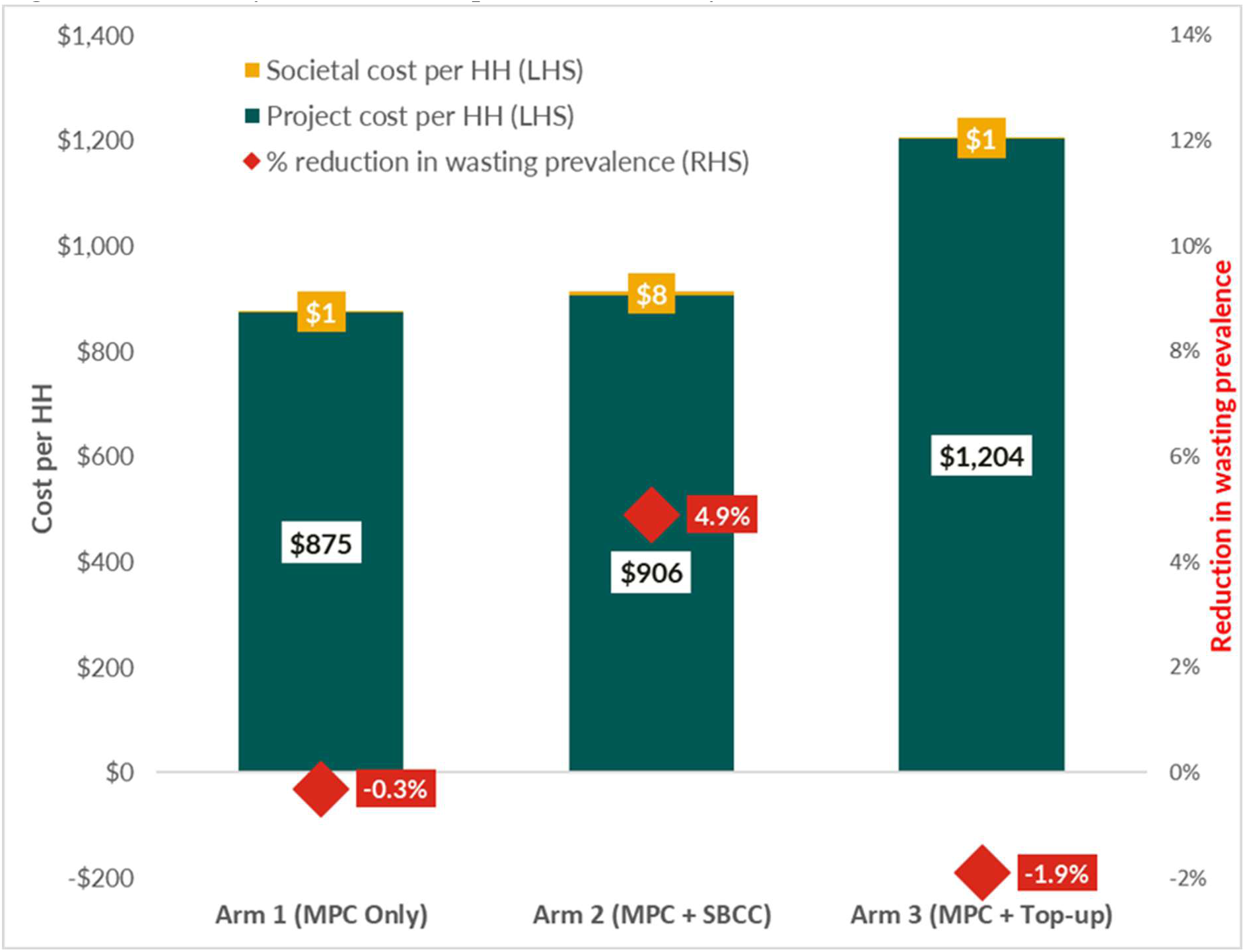
Cost Analysis Results Compared Across Study Arms.

By endline, there was no statistically significant difference in wasting prevalence change between Arm 3 and Arm 1, and additional costs spent on ineffective interventions are inherently not cost effective. Therefore, our overall assessment posits that Arm 2 was cost-effective compared to Arm 3. While Arm 2 had the greatest societal cost, requiring a beneficiary time investment of $8 per household, this investment only accounted for 0.91% of the total cost per household.

### Immediate Determinants

**Table 4** and Figure 4 summarize key household, child, and maternal factors associated with child malnutrition. At the immediate level, child diet diversity—including the intake of animal-source proteins—improved across all study arms, with Arm 3 showing the highest levels by endline. Reports of child illness declined in Arms 1 and 2 but increased in Arm 3.

**Figure 4.**
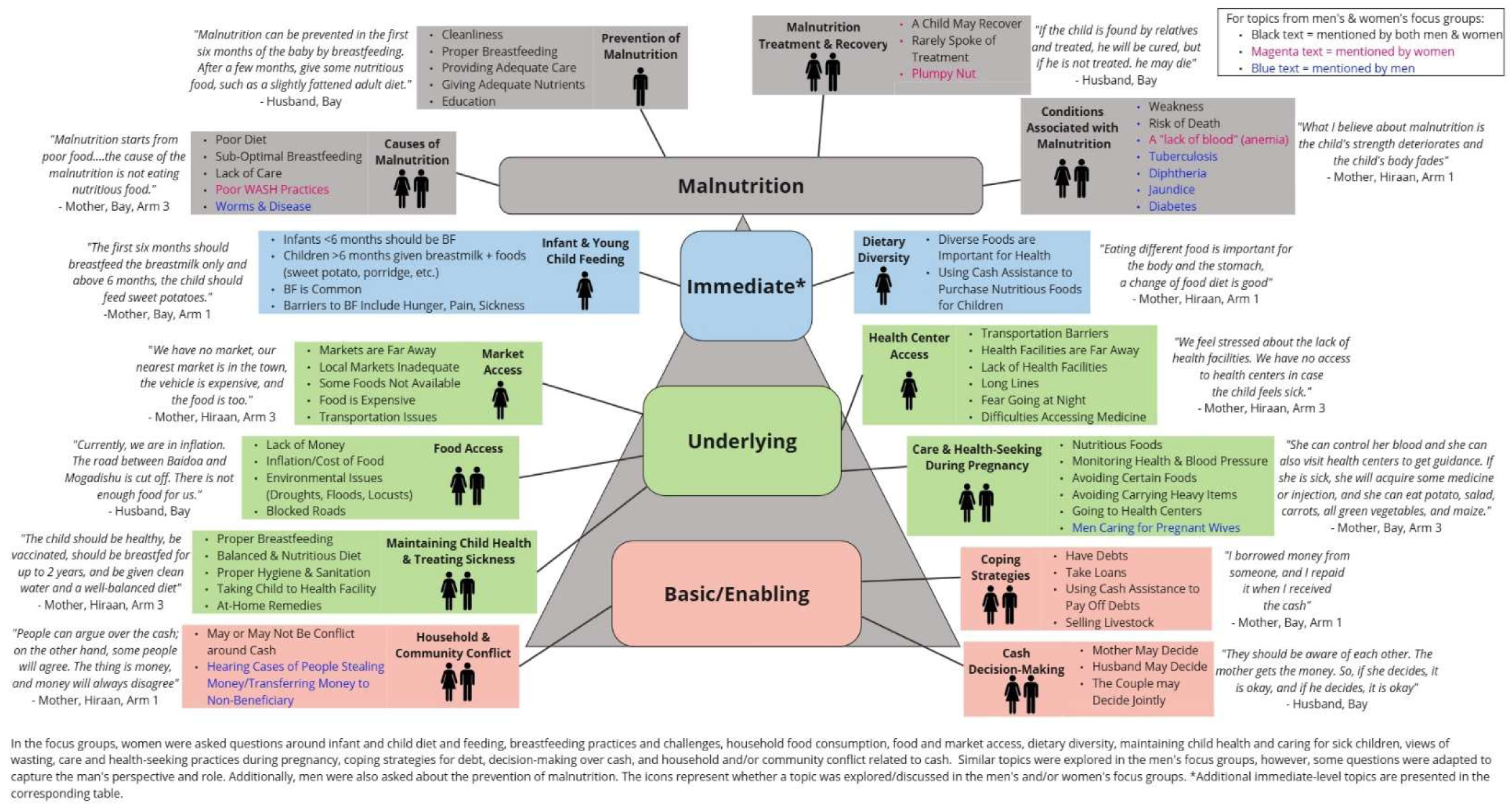
Qualitative Results.

**Table 4.** Household, Maternal and Child Factors related to Child Wasting by Study Arm Over Time.

| Child Wasting Drivers | Study Arm | Baseline Percentage (95% CI) | Midline Percentage (95% CI) | Endline Percentage (95% CI) |
| --- | --- | --- | --- | --- |
| **Immediate factors** |  |  |  |  |
| Meet Minimum Dietary Diversity for child (MDD-C), % |  | **n=415** | **n=904** | **n=1,022** |
|  | Arm 1 | 5.1 (2.6, 9.9) | 35.0 (29.1, 40.2) | 28.1 (23.3, 33.3) |
|  | Arm 2 | 12.0 (7.5, 18.8) | 33.4 (28.4, 38.9) | 32.3 (27.6, 37.1) |
|  | Arm 3 | 10.4 (6.1, 17.2) | 36.6 (31.4, 42.2) | 42.5 (37.3, 47.8) |
| Child received animal-sourced protein (meat, eggs, milk), % |  | **n=478** | **n=1,112** | **n=1,153** |
|  | Arm 1 | 53.1 (45.7, 60.4) | 76.2 (71.5, 80.4) | 73.5 (68.7, 77.8) |
|  | Arm 2 | 58.8 (50.9, 66.2) | 74.9 (70.2, 79.0) | 73.3 (68.7, 77.4) |
|  | Arm 3 | 56.6 (48.3, 64.6) | 69.4 (64.6, 73.8) | 81.6 (77.4, 85.1) |
| Had childhood illness* 2 weeks prior (malaria, diarrhea and/or cough), % |  | **n=477** | **n=1,112** | **n=1,153** |
|  | Arm 1 | 41.1 (34.0, 48.6) | 31.5 (26.8, 36.6) | 25.4 (21.2, 30.2) |
|  | Arm 2 | 31.4 (24.7, 39.1) | 31.8 (27.2, 36.7) | 27.8 (23.6, 32.4) |
|  | Arm 3 | 25.9 (19.3, 33.7) | 30.8 (26.4, 35.6) | 36.6 (31.9, 41.5) |
| **Underlying household factors** |  |  |  |  |
| Moderate-Severe Household Hunger Scale, % |  | **n=1,490** | **n=1,101** | **n=1,011** |
|  | Arm 1 | 48.1 (44.3, 51.9) | 32.8 (28.4, 37.5) | 44.6 (39.6, 49.8) |
|  | Arm 2 | 38.7 (35.1, 43.5) | 27.2 (23.2, 31.6) | 50.4 (45.5, 55.3) |
|  | Arm 3 | 35.5 (31.7, 39.5) | 18.5 (15.0, 22.5) | 43.7 (38.9, 48.7) |
| Borderline-Poor Food Consumption Score, % |  | **n=1,490** | **n=1,101** | **n=1,011** |
|  | Arm 1 | 68.0 (64.4, 71.4) | 32.5 (28.1, 37.2) | 52.9 (47.7, 58.0) |
|  | Arm 2 | 57.0 (53.1, 60.7) | 27.6 (23.6, 32.1) | 47.1 (42.3, 52.0) |
|  | Arm 3 | 50.8 (46.7, 54.9) | 34.5 (30.1, 39.2) | 38.6 (33.9, 43.6) |
| Reduced Coping Strategies Index Score (mean, CI) |  | **n=1,490** | **n=1,101** | **n=1,011** |
|  | Arm 1 | 14.5 (13.7, 15.3) | 13.5 (12.5, 14.5) | 14.5 (13.5, 15.5) |
|  | Arm 2 | 15.3 (14.5, 16.0) | 14.3 (13.2, 15.3) | 14.6 (13.5, 15.7) |
|  | Arm 3 | 14.9 (14.0, 15.7) | 12.4 (11.5, 13.3) | 14.1 (13.1, 15.0) |
| Received any childhood vaccination, % |  | **n=1,477** | **n=1,112** | **n=1,153** |
|  | Arm 1 | 50.9 (43.4, 58.2) | 62.7 (57.5, 67.7) | 79.8 (75.4, 83.7) |
|  | Arm 2 | 48.4 (40.7, 56.2) | 59.4 (54.3, 64.2) | 86.8 (83.0, 89.7) |
|  | Arm 3 | 46.2 (38.1, 54.4) | 67.6 (62.8, 72.1) | 75.4 (70.9, 79.5) |
| Crowded household ($\geq 5$ household members), % | | **n=1,490** | **n=1,261** | **n=1,153** |
|  | Arm 1 | 67.7 (63.5, 71.6) | 57.6 (52.7, 62.3) | 100 (-) |
|  | Arm 2 | 65.5 (61.2, 69.5) | 59.9 (55.1, 64.4) | 99.5 (98.0, 99.9) |
|  | Arm 3 | 55.1 (50.6, 59.5) | 39.3 (34.7, 44.1) | 100 (-) |
| Households practicing open defecation, % |  | **n=1,485** | **n=1,086** | **n=1,140** |
|  | Arm 1 | 23.5 (20.0, 27.4) | 28.7 (24.1, 33.8) | 22.7 (18.7, 27.3) |
|  | Arm 2 | 13.7 (10.9, 17.0) | 15.0 (11.7, 19.0) | 19.0 (15.4, 23.2) |
|  | Arm 3 | 21.3 (17.8, 25.2) | 27.3 (23.1, 32.0) | 33.5 (29.0, 38.4) |
| Households using unprotected drinking water source, % |  | **n=1,483** | **n=1,107** | **n=1,148** |
|  | Arm 1 | 41.1 (36.9, 45.5) | 45.1 (39.9, 50.4) | 30.0 (25.5, 35.0) |
|  | Arm 2 | 32.3 (28.4, 36.6) | 34.1 (29.5, 39.1) | 24.3 (20.3, 28.8) |
|  | Arm 3 | 43.5 (39.1, 48.1) | 31.0 (26.6, 35.8) | 27.2 (23.0, 31.9) |
| No access to handwashing facilities % |  | **n=1,489** | **n=1,112** | **n=1,153** |
|  | Arm 1 | 95.5 (93.3, 97.0) | 97.7 (95.5, 98.9) | 78.8 (74.2, 82.7) |
|  | Arm 2 | 99.0 (97.6, 99.6) | 94.4 (91.5, 96.3) | 83.3 (79.3, 86.6) |
|  | Arm 3 | 98.7 (97.2, 99.4) | 99.2 (97.6, 99.8) | 89.8 (86.3, 92.4) |
| Correct breastfeeding Initiation knowledge (within 1 hour), % |  |  | **n=1,096** | **n=1,148** |
|  | Arm 1 | N/A | 88.9 (85.0, 91.8) | 98.6 (96.7, 99.4) |
|  | Arm 2 | N/A | 93.5 (90.4, 95.6) | 99.5 (98.0, 99.9) |
|  | Arm 3 | N/A | 96.6 (94.3, 98.0) | 96.6 (94.3, 98.0) |
| Correct breastfeeding Initiation practice (within 1 hour), % |  |  | **n=1,095** | **n=1,148** |
|  | Arm 1 | N/A | 89.2 (85.44, 92.1) | 98.9 (97.1, 99.6) |
|  | Arm 2 | N/A | 94.2 (91.3, 96.2) | 99.3 (97.6, 99.8) |
|  | Arm 3 | N/A | 96.9 (94.6, 98.2) | 98.2 (96.3, 99.1) |
| Exclusive breastfeeding correct knowledge, % |  |  | **n=1,112** | **n=1,153** |
|  | Arm 1 | N/A | 56.2 (50.9, 61.3) | 70.2 (6.2, 74.7) |
|  | Arm 2 | N/A | 63.4 (58.3, 68.1) | 77.0 (72.6, 80.9) |
|  | Arm 3 | N/A | 68.9 (64.1, 73.3) | 68.5 (63.8, 73.0) |
| Exclusive breastfeeding_correct practice, % |  |  | **n=1,112** | **n=1,153** |
|  | Arm 1 | N/A | 57.6 (52.3, 62.7) | 72.0 (67.2, 76.5) |
|  | Arm 2 | N/A | 63.1 (58.1, 67.9) | 80.3 (76.0, 83.9) |
|  | Arm 3 | N/A | 69.7 (64.9, 74.0) | 72.9 (68.2, 77.1) |
| Optimal age for solid foods initiation_knowledge (6-8 months), % |  |  | **n=1,261** | **n=1,153** |
|  | Arm 1 | N/A | 56.9 (52.0, 61.6) | 35.9 (31.1, 41.0) |
|  | Arm 2 | N/A | 59.9 (55.2, 64.4) | 59.8 (54.9, 64.5) |
|  | Arm 3 | N/A | 58.3 (53.5, 62.9) | 44.0 (39.1, 50.0) |
| Optimal age for solid foods initiation _practice (6-8 months), % |  |  | **n=1,261** | **n=1,153** |
|  | Arm 1 | N/A | 31.6 (27.3, 36.3) | 38.7 (33.8, 43.8) |
|  | Arm 2 | N/A | 35.6 (31.2, 40.2) | 58.8 (53.8, 63.5) |
|  | Arm 3 | N/A | 35.5 (31.0, 40.2) | 45.3 (40.4, 50.2) |
| Household has $\geq 2$ U5 children, % | | **n=1,490** | **n=1,116** | **n=1,153** |
|  | Arm 1 | 31.3 (27.4, 35.5) | 17.7 (14.0, 22.0) | 29.8 (25.3, 34.8) |
|  | Arm 2 | 27.6 (23.9, 31.7) | 14.6 (11.4, 18.6) | 26.3 (22.2, 30.8) |
|  | Arm 3 | 21.0 (17.5, 24.9) | 13.1 (10.1, 16.9) | 18.4 (14.9, 22.6) |
| **Basic factors** |  |  |  |  |
| Total monthly household expenditure (mean \$, CI) | | **n=1,477** | **n=1,109** | **n=1,130** |
|  | Arm 1 | 113.1 (108, 118.0) | 158.3 (151.2, 165.4) | 155.9 (148.5, 163.2) |
|  | Arm 2 | 111.6 (105.5, 117.7) | 155.5 (148.9, 162.0) | 167.2 (161.6, 172.7) |
|  | Arm 3 | 95.0 (90.2, 99.8) | 186.4 (178.9, 193.9) | 106.8 (103.7, 109.9) |
| Average percent of monthly household expenditure on food (mean %, CI) |  | **n=1,477** | **n=1,109** | **n=1,130** |
|  | Arm 1 | 65.7 (63.9, 67.5) | 48.1 (46.2, 49.9) | 54.1 (52.3, 55.9) |
|  | Arm 2 | 70.6 (68.9, 72.2) | 51.4 (49.7, 53.1) | 55.7 (53.9, 57.4) |
|  | Arm 3 | 68.8 (67.2, 70.4) | 50.5 (48.9, 52.0) | 49.6 (47.9, 51.3) |
| Mother has no formal education, % |  | **n=1,489** | N/A | **n=1,149** |
|  | Arm 1 | 82.2 (78.6, 85.3) | N/A | 72.9 (68.0, 77.2) |
|  | Arm 2 | 82.6 (79.0, 85.7) | N/A | 67.9 (63.2, 72.3) |
|  | Arm 3 | 86.9 (83.5, 89.6) | N/A | 68.4 (63.6, 72.8) |
| Decision making on income is by mother or jointly, % |  | **n=1,475** | **n=1,108** | **n=1,148** |
|  | Arm 1 | 64.4 (60.1, 68.5) | 71.3 (66.3, 75.8) | 72.6 (67.7, 76.9) |
|  | Arm 2 | 64.4 (60.1, 68.5) | 76.5 (72.0, 80.6) | 76.9 (72.5, 80.1) |
|  | Arm 3 | 63.8 (59.3, 68.0) | 78.4 (74.0, 82.2) | 76.5 (72.1, 80.5) |
| Decision making on healthcare is by mother or jointly, % |  |  |  |  |
|  | Arm 1 | 68.6 (64.5, 72.5) | 71.3 (66.3, 75.8) | 81.5 (77.1, 85.2) |
|  | Arm 2 | 74.9 (70.9, 78.5) | 71.5 (66.7, 75.9) | 79.9 (75.7, 83.6) |
|  | Arm 3 | 73.3 (69.1, 77.2) | 69.9 (65.2, 74.3) | 80.2 (75.9, 83.8) |

These quantitative findings on diet and illness are reinforced by qualitative data from focus group discussions (FGDs). Across all arms, mothers demonstrated a strong understanding of the importance of dietary diversity for child health and nutrition, with many reporting that they used cash assistance to purchase more nutritious foods.

> *“It is used to buy nutritious food for the children, such as bananas, eggs, and soft food that will be of great help to them.”* – Mother, Hiraan, Arm 1
>
> *“Eating different food is important for the body and the stomach. A change of diet is good—if you keep on eating one kind of food, you won’t get nutrition from it.”* – Mother, Hiraan, Arm 1

Community members in both Bay and Hiraan also expressed a clear understanding of the causes of malnutrition and child illness. They emphasized the importance of a nutritious diet and breastfeeding, while acknowledging persistent barriers such as food insecurity and limited access to health services.

> *“Malnutrition can be prevented in the first six months by breastfeeding”* Husband, Bay, Arm 2
>
> *“Malnutrition results from weakness in the body… caused by lack of proper feeding, discontinuation of breastfeeding, and drinking unsafe water.”* – Mother, Hiraan, Arm 3

Yet, there are barriers to feeding the child a diverse diet and concerns about child disease contributing to malnutrition. Mothers in all arms were concerned about measles, diarrhea, and acute watery diarrhea (AWD) as major threats. A mother from Hiran (Arm 2) reported,

> *“This year there were more diseases such as measles and AWD… there is a shortage of vaccines for measles.”Underlying Determinants*

**Table 4** also summarizes food insecurity indicators—moderate/severe hunger and borderline/poor food consumption scores— improved in all arms from baseline to midline but rebounded somewhat by endline. Vaccination coverage increased across all arms, with Arm 2 achieving the highest rates by endline.

Access to protected water sources improved, though open defecation persisted and even increased in Arm 3. Handwashing facilities became more available, but gains were limited in Arm 3. Breastfeeding practices remained strong, with early initiation rates above 90% across arms. Knowledge and practice of exclusive breastfeeding exceeded 50% overall, rising most in Arm 2 (up to 80% at endline).

Qualitative data, presented in **Figure 4** from focus group discussions (FGDs) with mothers and fathers of children under five echoed these trends and contextualized the barriers to improving household food security. Participants frequently cited inflation, limited purchasing power, and the high cost of food as major challenges. Environmental shocks, such as drought, flooding, and locust infestations, as well as blocked roads and supply disruptions, were also mentioned as key constraints.

> *“The cash money is too small. When we go out to look for cereals, it is too expensive, and the farms we used to depend on were destroyed by locusts.”* – Mother, Hiraan, Arm 1

Monitoring of nutritious food in local markets was conducted by SC at baseline, midline and endline; data from market monitoring confirmed that food availability varied significantly by region and season, due to both typical supply patterns and access challenges Market monitoring methods are described in **Appendix S6** and the full market monitoring report may be made available on request from the corresponding author. In Bay region, cowpeas were unavailable in September (midline), groundnuts were missing in both September and December (endline), and spinach was absent in December. In Hiraan, availability gaps were more extensive: camel meat, eggs, cowpeas, canned oats, swiss chard, groundnuts, white maize, lentils, spinach, and pumpkin were consistently unavailable in Mataban and Mahas districts. However, Beletweyne District maintained availability of all tracked commodities.

FGD participants reflected this variability—some mothers noted market shortages, while others emphasized that food was available but unaffordable.

> *“There is a chance we do not have a suitable market where we can find everything we need.”* – Mother, Hiraan, Arm 2
>
> *“There are no big challenges. If you have your money, in the market you can peacefully buy anything.”* – Mother, Bay, Arm 3

Although some food prices remained stable or decreased, the cost of key nutritious and staple foods rose notably in both regions over the study period. In Bay, prices increased for vegetable oil, beans, maize, sorghum, banana, swiss chard, pumpkin, spinach, potatoes, and goat liver. In Hiraan, beans, lentils, sorghum, white rice, goat liver, lime, and onions saw similar inflation. These price hikes reinforced food access barriers voiced in the FGDs.

> *“When the price of food was normal, the money was enough. But when the price increased, we could only get half of rice and flour, and some oil and milk. One liter of oil is $7, and half the rice is about $30. It only lasts 15 days, then we have nothing.”* – Mother, Bay, Arm 3

Contributing factors identified through market monitoring included seasonal variability, flash flooding in Bay, drought in Hiraan, damaged supply routes, crop failure, increased taxes, and political instability, all of which were echoed by community members.

> *“Rice and spaghetti are mostly used, but our country’s food has not been harvested. The locusts have destroyed [the crops].”* – Mother, Hiraan, Arm 1
>
> *“Currently, we are in inflation. The road between Baidoa and Mogadishu is cut off. There is not enough food for us.”* – Husband, Bay, Arm 3

### Basic Determinants

In Table 4 highlights how household expenditures rose from baseline to midline, stabilizing in Arms 1 and 2 (∼$160), but dropping sharply in Arm 3 ($186 to $107). Spending on food declined from ∼70% to ∼50% across arms. Maternal education and household decision-making improved over time with minimal differences between arms.

These FGDs reinforce that the cash assistance was used for food, water, medical needs, school expenses, and other household needs. A few participants used cash in resourcefulways, such as for entrepreneurial endeavors.

> *“That cash assistance has been covering some of our basic needs like water expenses, electricity, educational fees, food, and milk for the children, and businessmen and women got revenue and profits from it.”* – Mother, Hiraan, Arm 2

Women in Arm 2 share their views on the benefits of SBCC explaining how they learned key skills and lessons related to nutrition and complimentary feeding.

> “*Earlier on I used to give only water and sugar but now I learnt exclusively breastfeeding in the first 6 months.”* – Mother, Bay, Arm 2*“The people got the messages, they bought nutritious food for their children, and the change was visible””* – Mother, Hiraan, Arm 2

## Discussion

### Summary

This cluster-randomized controlled trial provides robust evidence on the effectiveness and cost-effectiveness of a “CashPlus SBCC” intervention in reducing acute malnutrition among children under five in Somalia. Specifically, the addition of social and behavior change communication to unconditional cash transfers (Arm 2) led to a significant reduction in child wasting compared to UCT alone (Arm 1) and UCT plus a top-up cash amount (Arm 3). Arm 2 showed a 5.9 percentage point reduction and a 39.3% relative reduction in child wasting from baseline to midline and sustained improvements at endline, while Arm 3 – despite higher financial input – did not yield better nutritional outcomes. Moreover, Arm 2 was the most cost-effective strategy, achieving significant health impacts at a modest additional cost.

These findings are critical in the Somali context, where humanitarian resources are limited, and acute malnutrition remains prevalent. Our results underscore the importance of integrating SBCC with unconditional cash interventions to enhance their impact on nutrition, rather than relying on cash alone or increasing cash amounts without complementary education or support. We propose potential mechanisms of impact as guided by our findings in **Box 1**.

#### Box 1. Potential Mechanisms of Impact

The superior performance of the cash plus SBCC arm is likely explained by multiple reinforcing mechanisms. Qualitative data reveal that SBCC activities—such as mother-to-mother support groups and cooking demonstrations—enhanced caregivers’ knowledge of optimal infant and young child feeding practices. This knowledge translated into improved dietary diversity, especially in animal-source protein consumption, and better feeding behaviors. Furthermore, Arm 2 participants reported stronger awareness of health risks, hygiene practices, and early initiation of breastfeeding. Arm 2 also achieved 80% knowledge and practice of exclusive breastfeeding which reflects the success of the integrated SBCC approach and also signals strong potential for long-term improvements in infant and young child feeding (IYCF) behaviors and malnutrition prevention in the community. Improved vaccination rates and reduced illness episodes among children in Arm 2 also suggest that SBCC influenced health-seeking behaviors and broader care practices. In contrast, the additional cash in Arm 3 did not consistently improve nutrition outcomes. While dietary diversity was highest in this group by endline, the lack of accompanying SBCC may have limited the efficient and nutrition-sensitive use of cash. Some households in Arm 3 faced higher rates of open defecation and worsening food consumption scores, highlighting that additional cash cannot compensate for deficiencies in health infrastructure or knowledge.

Market monitoring and qualitative data illustrate how the broader food system moderated intervention impacts. Food availability varied across regions and seasons, with key nutritious items like spinach and groundnuts intermittently unavailable, particularly in Hiran. Participants also cited food inflation, environmental shocks (flooding, locusts), and transport disruptions as constraints on their purchasing power and dietary quality, particularly after midline. These environmental shocks may explain the partial erosion of earlier gains and underscore the need for resilience-building components in humanitarian programming.

### Comparison with Previous Studies

Our findings contribute to a growing but still limited body of rigorous evidence on the nutritional impacts of cash-based interventions in humanitarian settings (**Appendix 12**). While previous studies have demonstrated that cash transfers can improve food security and dietary diversity [63–72], few have measured direct impacts on acute malnutrition using anthropometric indicators such as WHZ. Our results confirm the potential of cash transfers to positively affect nutrition outcomes, particularly when combined with SBCC – a result consistent with a quasi-experimental study in Somalia that found positive impacts of cash plus nutrition counseling.[73] However, that study lacked a cash-only comparison group, limiting its ability to attribute effects specifically to SBCC.

Our findings also diverge from studies suggesting that higher-value cash transfers can yield greater impacts on nutrition.[74–76] In our context, increasing the cash amount alone (Arm 3) did not yield better outcomes and may have introduced complexity or unintended spending behaviors that diluted nutritional benefits (e.g. on debt reduction or social activities). This highlights the importance of context-specific program design and the potential diminishing returns of increasing transfer size without behavioral support.

Our findings contrast with studies suggesting that higher cash transfers inherently lead to better nutrition outcomes by showing that Arm 2 – cash plus SBCC – was both more effective and far more cost-efficient than a higher-value cash transfer alone. This challenges the assumption that “more cash equals more impact” and highlights the diminishing returns of increasing transfer size without addressing behavioral and market constraints. [74,75,77,78] In contrast, the modest cost of SBCC in Arm 2 produced strong returns – even when factoring in opportunity costs— echoing findings from other evaluations where layering nutrition counseling or SBCC onto cash transfers significantly improved program efficiency and outcomes.[77–79]

Our findings add nuance to the existing evidence base on the duration of cash assistance and its impact on nutrition outcomes. They underscore the importance of considering program duration when evaluating the impact of cash assistance. While short-term transfers can provide immediate relief, they may not allow households to make strategic, longer-term decisions around spending, savings, or investments in nutrition and health. More research is needed to assess how households adjust their financial decisions when cash support is sustained over a longer and more predictable timeframe. While several studies have shown that longer-duration or sustained cash transfers lead to stronger improvements in nutrition [74,75,80], our trial observed the greatest reduction in child wasting at 3 months, particularly in the cash + SBCC arm, with limited additional gains by 6 months. This plateau effect diverges from some literature and may reflect contextual challenges in Somalia. Between midline and endline, the study regions experienced significant flooding, price inflation, and market access disruptions [36], among other challenges, all of which likely constrained households’ ability to maintain (or make further gains) improved diets and care practices. These environmental and economic shocks may have masked the potential for further benefit from continued cash assistance. This underscores the importance of embedding adaptive, tailored to the target groups and context-sensitive approaches in program design and continued research, especially when operating in volatile humanitarian settings.

### Strengths

This study offers several methodological and practical strengths that advance the evidence base on cash-based interventions for nutrition in humanitarian settings. First, it employed a rigorous cluster-randomized controlled trial design across multiple sites, allowing for strong causal inference – an improvement over the quasi-experimental designs that dominate this field. Second, the inclusion of three intervention arms (cash only, cash plus SBCC, and cash plus top-up) enabled direct comparison of additive components, helping to isolate the value of behavior change communication. Third, the study integrated both incremental cost-effectiveness analysis and societal cost assessment, providing rare but essential insights into value for money and opportunity costs – critical for donor decision-making; strengths of our approach are further details in **Appendix 13**. Fourth, outcomes were measured using gold-standard anthropometric indicators such as WHZ and child wasting prevalence and incidence, enhancing comparability with global nutrition targets. Fifth, the study’s real-time market and qualitative monitoring allowed contextual interpretation of findings in light of environmental shocks like flooding and inflation, bolstering external validity. Sixth, the SBCC component was designed to be scalable, low-cost, and embedded within existing community structures, offering a practical and replicable model for humanitarian actors seeking effective, locally adaptable solutions. Further research is needed, however, to determine which specific components of SBCC yielded the greatest impact. A key strength of this study was its robust analytical approach, which included both intention-to-treat and per-protocol analyses, as well as assessments of wasting incidence and prevalence.

These complementary methods confirmed the consistency of the findings and strengthened confidence in the internal validity of the observed intervention effects. Finally, geographic coverage across both Bay and Hiraan regions strengthens the generalizability of the findings within diverse humanitarian contexts in Somalia.

### Limitations

Several limitations of this analysis exist. While attrition was moderate (∼22%), it was not systematically different across arms or associated with baseline nutritional status, reducing potential bias. The intervention was implemented in a dynamic and highly fragile context, with significant flooding, inflation, and supply chain disruptions occurring between midline and endline. These contextual shocks may have constrained household purchasing power and food access, potentially attenuating the nutritional impacts observed in later months. Nonetheless, the study’s adaptive design – featuring ongoing market monitoring, qualitative assessments, and responsive implementation – helped mitigate some of these challenges by allowing for real-time interpretation and programmatic adjustments. Another limitation is that, although maternal nutrition and health behaviors were measured, the study was not powered to detect statistically significant changes in maternal nutrition outcomes; further research on this is needed. A key limitation of this study is the absence of a study arm that assessed the combined impact of increased cash (top-up) and social and SBCC. As a result, we are unable to determine the potential synergistic effects of this combination on nutrition outcomes. While this would have been a valuable comparison, resource constraints limited the number of intervention arms we could feasibly implement within the scope of the study. Additionally, variation in SBCC exposure across clusters and potential spillover effects could not be fully accounted for and may have influenced outcomes. Finally, despite rigorous market monitoring, participants were asked to recall market prices and availability for September, posing a risk of recall bias. Though our cost analysis was robust, limitations such as subjectivity in opportunity costs and available secondary data existed (**Appendix 13**).

### Policy and Programmatic Implications

Our findings have clear implications for humanitarian policy and programming. First, they provide compelling evidence that cash alone is not enough to improve child nutrition in crisis settings. Behavioral components are essential for converting economic inputs into nutritional gains. Next, the study supports investments in cost-effective, scalable SBCC models and platforms—such as mother-to-mother groups – which can be layered onto existing cash transfer mechanisms with minimal additional cost [81]. Also, donors and implementers should recognize that increasing cash transfer size may not yield proportional improvements in nutrition and may, in some cases, divert funds from more effective, behaviorally informed approaches. Particularly in contexts where markets are underdeveloped or volatile, adding cash without improving decision-making support can lead to suboptimal outcomes. Additionally, integrated monitoring of markets, dietary trends, and environmental shocks is essential to adapt programming dynamically. Currently, cash and nutrition programs are often implemented through siloed systems, with limited coordination between actors such as the Somali Cash Consortium and the Nutrition Cluster, as well as across different ministries. This fragmentation weakens the effectiveness of interventions and limits their potential for impact. A key policy recommendation emerging from this research and stakeholder discussions is to strengthen coordination by linking cash and nutrition actors through established platforms like the Scaling Up Nutrition (SUN) Movement and the Office of the Prime Minister. A dedicated intersectoral working group should be established to facilitate collaboration, align objectives, and improve the coherence of policy and programmatic responses. Finally, adaptive design, as employed in this trial, should be considered a best practice for future evaluations in complex emergencies.

### Conclusion

In conclusion, this study provides the strongest evidence to date from Somalia on the comparative effectiveness and cost-effectiveness of “CashPlusSBCC” programming to prevent child wasting. The addition of SBCC to cash transfers (Arm 2) significantly outperformed both cash alone and increased cash, offering both improved and sustained nutritional outcomes and high value for money. These findings make a compelling case for integrated, multi-component humanitarian assistance strategies that combine financial support with education, especially in fragile and food-insecure settings. As humanitarian funding becomes increasingly constrained, such evidence-based approaches are vital for ensuring that scarce resources deliver maximum impact for vulnerable populations.

### Ethics

All participants were informed of the study purpose, the use of the collected data, and any potential risks to participation. Individuals that agreed to participate signed an informed consent form. Approval for this study was obtained from the Johns Hopkins Bloomberg School of Public Health Institutional Review Board (protocol code24476), the Somalia Ministry of Health and Human Services (XAG/203/23), and the Ethics Review Committee at Save the Children International.

### Data Availability

This data was collected on a vulnerable population linked to a large-scale cash assistance program in Somalia. The data may be available upon request with permission from local study investigators.

### Funding

Funding for the research study was obtained from Elrha (grant number 200011671). Save the Children Somalia implemented the CashPlus for Nutrition Project, funded by the United States Agency for International Development/Bureau of Humanitarian Assistance.

### Authorship Contributions

Conceptualization of the original study design: NA,QK, MT, FL, SAM, MAN, MBM, DIJ, DG, MO, MO, AA, AAM, AA, SW; writing (original draft preparation) – NA, KKA, SG, SG, QK, EM, SW; writing (review and editing) – NA, KKA, SG, SG, QK, MT, FL, SAM, MAN, SA, EM, MO, MO, AA, AAM, AA, SW. All authors have read and agreed to the published version of the manuscript.

## Supporting information

Supplemental Results Tables and Figures

Supplemental Technical Appendix

## Acknowledgments

We would like to acknowledge the donors who provided generous funding for this research, the study participants and their families, the Ministries of Health in Somalia who supported the study, and the teams of dedicated staff who collected data.

## Conflicts of Interest

The authors declare no conflicts of interest.

## Notes

### Competing Interest Statement

The authors have declared no competing interest.

### Clinical Trial

NCT06642012

### Funding Statement

This study was funded by Elrha (grant number 200011671). Save the Children Somalia implemented the CashPlus for Nutrition Project, funded by the United States Agency for International Development/Bureau of Humanitarian Assistance.

### Author Declarations

The Johns Hopkins Bloomberg School of Public Health Institutional Review Board gave ethical approval for this work (protocol code: 24476). The Somalia Ministry of Health and Human Services gave ethical approval for this work (XAG/203/23). The Ethics Review Committee at Save the Children International gave ethical approval for this work.

