## Supplemental Technical Appendix for "The Effectiveness and Cost-Effectiveness of CashPlus Interventions to Prevent Acute Malnutrition in Somalia: Evidence from an Adaptive Cluster Randomized Control Trial"

**Technical Appendices: S1-S13**

**Table of Contents**

Appendix S1. Mobile Messages Received by BHA Program Participants

Appendix S2. Arm 2 SBCC Intervention Details

Appendix S3. Outcome Definitions for CU5 and Mothers

Appendix S4. Data Collection and Measurement Procedures

Appendix S5. Exposures: Household Survey Indicators and Definitions

Appendix S6. Data Analyses Methods

Appendix S7. Focus Group Discussion Data Collection & Analysis Details

Appendix S8. Names and Roles of Program Staff Consulted for Cost Analyses

Appendix S9. Cost Analysis Indicators

Appendix S10. Market Monitoring Data Collection Sites

Appendix S11. Food Commodities Monitored During Market Monitoring

Appendix S12. Effectiveness and Cost-Effectiveness of Cash-Based Interventions on Nutrition Outcomes

Appendix S13. Strengths and Limitations of Cost Analysis Approach

**Appendix S1. Mobile Messages Received by BHA Program Participants**

**English Translation**

Are you aware of the importance of good nutrition for your infants and young children?

We breastfeed our babies exclusively for 6 months because breastmilk is the ideal food for infants and provides all the energy and nutrients that the infant needs for the first months of life.

We breastfeed until our children are two years old because the first two years is our children's opportunity to grow and be ready for the world.

We breastfeed because breastmilk is safe, clean and contains antibodies that help protect against many common childhood illnesses.

Remember to practice good hygiene and sanitation to help prevent illness and ensure your child stays healthy.

Talk to your healthcare provider or community health worker to learn more about IYCF and how you can provide your child with the best possible start in life.

Together, we can give our children the gift of good health and a brighter future. Don't wait, start practicing good Infant and Young Child Feeding practices today.

**Appendix S2. Arm 2 SBCC Intervention Details**

SBCC sessions covered the following topics: breastfeeding within the first hour/importance of colostrum; exclusive breastfeeding for 6 months; attachment and positioning of the baby at the breast; breastfeeding misconceptions and myths; complementary feeding (continuing breastfeeding and giving other foods/liquids); medical care; hygiene; common conditions that can affect breastfeeding; physical and mental stimulation of the child; relationships (i) with partner/husband and (ii) with other family members, (e.g. mother-in-law); mother’s safety/fear/worries; nutrition of pregnant and lactating mothers.

Save the Children identified 11 staff members to serve as community-based IYCF promoters to train mothers in IYCF practices and engagement strategies for mother-to-mother (M2M) support groups (4 promoters in Bay and 7 in Hiran). SBCC M2M sessions were then led by trained lead mothers who facilitated discussions among 10-15 mothers. Sessions occurred weekly, covering one of the above topics each week (for a total of 12 sessions in 12 weeks), and group members decided on the day of the sessions. Sessions lasted from 45-60 minutes. The SBCC sessions were held in a safe place to enable mothers to exchange ideas, share experiences, give and receive information, and offer and receive support in breastfeeding, child-rearing, and women’s health. The lead mother also followed-up with and managed absenteeism during sessions.

In addition to the SBCC sessions, Save the Children organized cooking/food demonstrations for support groups to showcase locally available and affordable nutritious food.

Some mothers enrolled in Arm 2 missed some of the Mother-to-Mother Support Group (MtMSG) sessions, while others migrated due to search of pasture and water for their livestock. The implementation team provided make-up sessions for the mothers who missed some MtMSG sessions. For those who migrated, the implementation team attempted to identify their new locations and collaborated with the local community to arrange MtMSG sessions that were more accessible.

**Appendix S3. Outcome Definitions for CU5 and Mothers**

| ***Malnutrition Guidelines for CU5*** | |
| --- | --- |
| Without Acute Malnutrition | MUAC^1^ ≥ 125 mm and WHZ^2^ ≥ -2 SD,  without edema |
| With Moderate Acute Malnutrition (MAM) | 115 mm ≤ MUAC < 124.9 mm  and/or -3 SD ≤ WHZ < -2 SD,  without edema |
| With Severe Acute Malnutrition (SAM) | MUAC < 115mm,  and/or WHZ < -3 SD,  and/or presence of edema |
| ***Stunting Guidelines for CU5*** | |
| Without Stunting | HAZ^3^ > -2 SD |
| Moderate Stunting | -3 SD < HAZ ≤ -2SD |
| Severe Stunting | HAZ ≤ -3 SD |
| ***Malnutrition Guidelines for Mothers of CU5*** | |
| Overweight | MUAC > 300mm |
| Normal | MUAC ≥ 230 mm |
| With Moderate Acute Malnutrition (MAM) | MUAC < 230 mm |

^1^MUAC: Mid-upper arm circumference

^2^WHZ: Weight-for-height z-score

^3^HAZ: Height-for-age z-score

**Appendix S4. Data Collection and Measurement Procedures**

Training for enumerators covered standardized data collection procedures, anthropometric measurement techniques, and tool familiarization. To ensure consistency and data quality, the same team of enumerators was retained throughout the study period, with refresher training sessions conducted before each subsequent data collection phase. Data was collected electronically using KoboToolbox, a secure mobile data collection platform. At baseline, we collected anthropometric measurements for all children under 5 years (CU5) in the household and the mother. We also collected child-level information for the youngest U5 child in the household at baseline, who was then followed longitudinally throughout the study period, regardless of any new births in the household. The survey captured comprehensive household information including socioeconomic indicators, food security metrics, WASH practices, healthcare access, and other factors in alignment with the UNICEF Conceptual Framework on Maternal and Child Nutrition.

Standardized equipment was used for all anthropometric measurements. Height/length measurements were conducted using UNICEF measuring boards with a fixed base and adjustable headpiece, providing precision to 0.1 cm. Children under 24 months were measured in a recumbent position, while those 24 months and older were measured standing. Weight measurements were obtained using SECA 874 battery-operated digital scales, accurate to 0.1 kg. For infants and young children unable to stand independently, weight was measured using the tare function while being held by their mothers. Mid-upper arm circumference (MUAC) was measured using standard non-stretch MUAC tapes with 0.1 cm precision. Quality control measures included daily field supervision and real-time data quality checks. The electronic data collection forms incorporated built-in validation rules to minimize data entry errors.

**Appendix S5. Exposures: Household Survey Indicators and Definitions**

| **Enabling Causes** | |
| --- | --- |
| *Context, Capital, and Resources* | |
| Demographic Information and Household Characteristics | Participants were surveyed on a variety of household and demographic characteristics including:   - Age of the mother - Sex of the Head of Household - Pregnancy Status of the Mother - Number of CU5 in the household - Displacement due to flooding (endline) - Receipt of Cash Assistance/Livelihood Support from other programs     Nutrition sensitive/cash programs (cash assistance, food baskets, livelihoods support |
| Maternal Education | Mothers were asked what their highest level of education was:   - No formal education - Madrasa - Primary - Secondary |
| Maternal Empowerment and Decision-making | Mothers were asked about their involvement in household decision-making in three areas:   - Income - Healthcare - Purchases   For each area, the decision-maker was either:   - Father - Mother - Joint decision-making |
| Household Assets: Principal Component Analysis | Household assets and wealth were evaluated using the Principal Component Analysis index. The index uses asset indicators rather than income data to visualize long-term economic status.    The asset index was constructed from these assets and housing characteristics:   - Livestock ownership - Land ownership (in hectares) - Back account ownership - Household items (electricity, radio, TV, telephone, computer, refrigerator, internet air conditioning) - HH Assets (watch, mobile phone, bicycle, scooter, donkey cart, truck, canoe, tractor, ox plough) - Crowding status - Floor - Roof - Walls - Type of toilet - Water Source   Household were then categorized into asset index quintiles: poorest, poorer, middle, richer, and richest. |
| Household Expenditures | Participants were surveyed on monthly expenditures (absolute dollar value) and expenditure categories.    Expenditure categories included: food, water, medical needs, agriculture, hygiene, electricity, maternal/child health needs, social activities, transport, communication, clothes and NFIs, debt repayment, fuel, rent, school fees, and savings. |
| **Underlying Causes** | |
| *Household Food Security* | |
| Household Hunger Scale | The Household Hunger Scale (HHS) measures the scale of households’ food deprivation.    Method: Participants are asked if their HH has experienced any of the following in the past 30 days and how often?   - No food to eat of any kind because of lack of resources to get food. - A HH member had to sleep hungry because there was not enough food. - A HH member had to go a whole day and night without eating anything at all because there was not enough food.     Frequencies are coded as rarely (1-2 times), sometimes (3-10 times), and often (>10 times).    Scores are summed and categorized as:  0-1: Little to no hunger  2-3: Moderate hunger  4-6: Severe hunger |
| Food Consumption Score | Measures how often household consume food items from the different food groups during a 7-day reference period.    Method: The consumption frequency of each food group is multiplied by its weight.    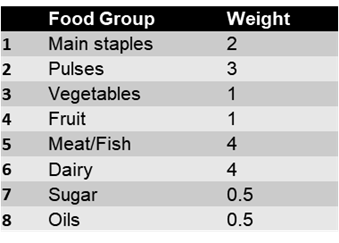    Scores are summed and categorized as:  0-28: Poor  28-5-42: Borderline  >42: Acceptable    Alternative thresholds for FCS categories were used due to high oil and sugar consumption in Somalia. |
| Reduced Coping Strategies Index | Measures the frequency and severity of HH food consumption behaviors due to food shortage in the 7 days prior.    Method: Participants are asked a series of 5 weighted questions and how often they have experienced these in the past 7 days.    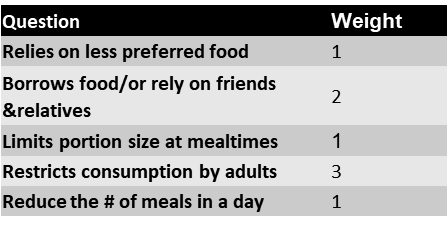    Scores are summed and range from 0-56. Higher scores mean households increasingly use worse coping mechanisms and have increased food insecurity. |
| *Unhealthy Household Environment* | |
| Household Crowding | Participants provided a household roster at survey timepoints.    Household crowding was defined as:  Not crowded: Less 5 people in the HH  Crowded: 5 or more people in the HH |
| Water, Sanitation, and Hygiene Practices (WaSH) | Participants were asked questions about their access to:   - Handwashing facilities with water and soap - Improved/unimproved sanitation facilities - Improved/unimproved water sources     Sanitation facilities and water sources were classified as improved or unimproved based on the DHS classification.    Sanitation:  Improved: flushing toilet, pit latrine, composting toilet  Unimproved: open defecation in a bush or field, bucket, other    Water Source:  Improved: piped water, public tap/standpipe, protected well, protected spring, rainwater, tanker truck, cart with small tank  Unimproved: unprotected well, unprotected spring, surface water, or other |
| *Inadequate Care and Feeding Practices* | |
| Maternal and Child Health | Participants were surveyed on topics related to maternal and child health including place of last birth, whether their last birth was assisted by a skilled-birthing attendant, caesarean birth, and child vaccination history. |
| Malnutrition Screening | Participants were asked if they or their child received malnutrition screening in a health facility, in the community, or at home in the last 3 months, whether they were referred for malnutrition treatment, and whether their child is currently enrolled in wasting treatment. |
| SBCC Indicators | Participants were surveyed on topics related to knowledge and practice of health and ICYF behaviors, including initiation and exclusivity of breastfeeding, initiation of liquid and solid foods, water treatment, critical handwashing moments, and awareness of M2M support groups (Arm 2 only). |
| **Immediate Causes** | |
| *Inadequate Dietary Intake* | |
| Minimum Dietary Diversity – Child | Measures how many of these food groups the child consumed the day before survey data collection:   - Breastmilk - Grains, roots, tubers - Legumes, nuts - Dairy products - Flesh foods (meat/fish) - Eggs - Vitamin A rich fruits and vegetables - Other fruits and vegetables     Scores range from 0 (consumed none of these food groups) to 8 (consumed all of these food groups) and dietary diversity is categorized as:   - Does not meet dietary diversity: <5 food groups - Meets dietary diversity: 5 or more food groups |
| *Diseases* | |
| Childhood Illness | Participants were asked whether their child experienced diarrhea, cough, or fever, within the past two weeks. |

**Appendix S6. Data Analysis Methods**

***Quantitative Data Management and Statistical Analysis***

Data quality checks and validation were conducted by data analysts in near real-time and verified by field teams. Baseline characteristics were summarized using descriptive statistics and we tested for differences across arms. Continuous variables were presented as means and standard deviations, while categorical variables were reported as frequencies and percentages. Main outcomes analyses were based on intent-to-treat prevalence (ITT) approach. We conducted several sensitivity analyses to assess the robustness of our findings, including per protocol (PP) analysis, comparison of prevalence vs incidence analysis, different approaches to handling missing data (complete case analysis vs available case analysis), and different specifications of the time variable (3 months vs 6 months). We also performed a separate analysis for participants lost to follow-up to assess differences from the main cohort. We used multilevel mixed-effects regression models for linear mean change in anthropometric indices and binary outcomes with 95% confidence intervals (CI) and presented results as crude, partially adjusted and fully adjusted for baseline factors for 3 arms at 3 months and 6 months.

We employed a difference-in-differences (DiD) approach using logistic regression to estimate the intervention effects. The DiD compared changes in child and maternal outcomes between intervention arms (arm 2, arm 3) and control arm (arm 1) from baseline to endline, with results expressed as odds ratios (ORs) and CI. We developed three model specifications: an unadjusted model examining only the treatment effect; a partially adjusted model controlling for cluster, child age, sex and wasting at baseline; and a fully adjusted model incorporating time-varying covariates such as maternal factors, household expenditures and WASH indicators. We computed post-estimation statistics including adjusted predictions at the means and average marginal effects for each study arm across time points. Panel effects was confirmed using the Breusch-Pagan Lagrange multiplier test. For comparing multiple intervention arms, we employed the Wald test to assess joint significance, while overall intervention effects were evaluated using likelihood ratio tests. To account for multiple comparisons across the three arms, we applied the Bonferroni correction to maintain the family-wise error rate at α = 0.05. All analyses maintained a significance level of p<0.05, and results are reported following CONSORT guidelines for cluster randomized trials. Data management and analysis were performed using STATA Version 18 (StataCorp, College Station, TX, USA).

***Qualitative Methods***

An explanatory sequential approach was taken, collecting qualitative data after the quantitative data in order to further explore and contextualize quantitative findings. Focus group discussions were held with mothers and fathers of CU5 beneficiaries of the BHA program, following a semi-structured guide exploring participants’ views on topics such as the BHA program implementation, the use of cash, dietary diversity and food security. A total of 6 focus groups with mothers (one per trial arm per region) and 2 focus groups with husbands (one per region, combining trial arms) were conducted, with 6-8 participants per focus group for a total of 48 mothers and 14 husbands. Focus groups were conducted, recorded, and transcribed in Somali and then translated into English for analysis. A mix of inductive and deductive approaches were undertaken for analysis. Additional details describing the focus group guide, data collection, and analysis are provided in **Appendix S7.**

***Cost Analysis Methods***

The cost efficiency and cost-effectiveness analyses in this study use a mixed-methods approach to triangulate findings from three retrospective data sources: 1) a desk review, 2) cost estimation and modelling, and 3) consultations with the SC Somalia country office research team.

Data were gathered from project financial reports and focus groups regarding all financial costs of delivering the intervention, including cost of financial procurement (i.e. cash, other SBCC activities, etc.) and implementation costs (i.e. cash delivery costs, training costs, staff salaries, etc.). Quantitative data was analyzed through a retrospective, top-down costing approach: activity-based costing and step-down cost accounting. Data were cleaned and processed, and then cost estimates were confirmed through group and individual consultations with program implementation staff and in-country cash and nutrition experts to include all direct, shared, and ‘societal’ costs of the intervention (list of staff consulted is available in **Appendix S8**). Costs of delivering the program activities and interventions were linked to the outcome of interest (CU5 wasting prevalence) to evaluate cost-effectiveness.

All costs and indicators were calculated in $USD. Cost-efficiency and societal cost analyses were compared across study arms and regions. Societal costs were calculated based on beneficiary time investment for participation in the study activities and program. Further details on the cost data, indicators used, and their definitions can be found in **Appendix S9**.

***Market Monitoring Methods***

Market monitoring focused on foods commonly consumed and typical of an energy-based diet as well as fresh and nutritious food items consumed by CU5 and PLW. A questionnaire was developed and administered by trained data collectors to 160 vendors in Bay (n=48) and Hiraan (n=112) in July 2023 and 111 vendors in Bay (n=33) and Hiraan (n=78) in December 2023. The questionnaire asked about the prices of commodities and reasons for changes in pricing. During the December 2023 data collection, recall data was also collected for September 2023 to have three timepoints (July, September, and December) in line with the timing and duration of the trial. Data collectors underwent a one-day training prior to data collection. In addition to interviewing vendors, data collectors also observed markets, took photos, and engaged with stakeholders to assess commodity availability, pricing, and factors affecting these characteristics. A list of the market sites is available in **Appendix S10**.

The price and availability of foods were reported by district (Baidoa in Bay region and Beledweyne, Mahas, and Mataban in Hiraan region), with foods being categorized into four main types of commodities for an energy-based diet: sentinel foods, fruits and vegetables, cereals, and protein sources. The list of specific food items monitored is provided in **Appendix S11**.

The lump sum value of commodities was calculated for each district. Changes in market prices over time were analyzed by comparing prices from the three data collection points (July, September, and December).

The market monitoring report is available by request from the corresponding author.

**Appendix S7. Focus Group Discussion Data Collection & Analysis Details**

Focus group discussions were held with mothers and fathers of CU5 beneficiaries of the BHA program. The midline survey dataset served as the sampling frame for selecting participants, and mothers were sampled based on education status, age, and head of household status, with the aim of sampling a diverse set of participants. Additionally, participants’ distance from their household to the FGD site was also considered for feasibility purposes. The men’s focus groups were comprised of the husbands of the mothers who were selected based on this sampling frame and criteria.

Mothers’ and husbands’ focus group discussion guides were created and explored the following major themes: the BHA program; the use of cash; health-seeking behaviors during pregnancy (or supporting healthy pregnancy for the husbands’ focus groups); child sickness and health-seeking behaviors when sick; and dietary diversity and food security. These guides are available in the supplementary materials of the related published study protocol.

A total of 6 mothers’ focus group discussions were conducted, with one focus group for each study arm in each region (Bay and Hiraan). One focus group per region was conducted with husbands, combining participants from all three trial arms, for a total of 2 focus groups with husbands. Each focus group was comprised of 6-8 participants, with a total of 48 mothers and 14 husbands participating.

Focus groups were conducted in central community locations in January 2024, about one month after the endline data was collected in December 2023. Trained enumerators confirmed enrollment criteria, obtained informed consent from each participant, and then used the relevant guide to facilitate a structured discussion. Focus groups were conducted, recorded, and transcribed in Somali and were then translated into English for analysis.

Focus group discussions were analyzed through a mix of inductive and deductive approaches following an iterative five-step analytic process. (25) As the focus groups followed a structured guide, the focus group discussion guides informed initial codes, and two study team members then utilized a two-phased approach to develop the codebook, with an initial round of rapid in vivo coding of a designated transcript, followed by a second round of coding to create additional codes. A structured codebook detailing each code was developed and shared with the qualitative data analysis team. (26) ATLAS.ti v. 24 software was used for coding, and additional codes were added during the coding process as necessary. Each transcript was coded by 2-3 team members, and versions of coded transcripts were merged to generate one final coded transcript for each focus group. Reports for each code were exported from Atlas. Both exploratory analysis and explanatory sequential analysis approaches were undertaken, using the qualitative data to attempt to contextualize and understand quantitative findings. (27) Qualitative findings were summarized and organized according to levels of the UNICEF conceptual framework, and data triangulation was achieved by comparing and contrasting findings from both the mothers’ and husbands’ focus groups with the quantitative findings. (28)

**Appendix S8. Names and roles of stakeholders and program staff consulted for cost analyses**

| **Name** | **Role** | **Organization** |
| --- | --- | --- |
| Adan Yusuf Mahdi | Senior Nutrition TA | SC Somalia |
| Abdullahi Arays | Head of REALM | SC Somalia |
| Said Gaafaa | Research Advisor | SC Somalia |
| Meftuh Omer | Head of Child Survival | SC Somalia |
| Muhamud Ali Nur | Area MEAL Manager | SC Somalia |
| Maimun Gure | Nutrition PM | SC Somalia |
| Dahir Gedi | Food Security and Livelihoods PM | SC Somalia |
| Sadiq Abdikadir | Area Research Coordinator | SC Somalia |
| Adam Faradhuub | Research Manager | SC Somalia |
| Mohamed Billow Mahat | Nutrition Focal Point | SC Somalia |
| Nimo Hussein Guleid | Nutrition Program Officer | SC Somalia |
| Simon Fuchs | Lead Economic Advisor | SC International |

**Appendix S9. Cost Analysis Indicators**

*Cost Efficiency*

To evaluate cost-efficiency of the intervention, three key indicators were used:

1) Cost Per Household: the total amount of resources spent per beneficiary HH over the program duration.

2) Cash Transfer Ratio (CTR): the amount of non-cash resources spent per $1 of cash transferred to HHs.

3) Operational Efficiency: the amount of resources spent to reach each beneficiary HH per month, irrespective of cash transfer amount.

*Cost-Effectiveness*

To evaluate cost-effectiveness of the intervention, one key indicator was used:

Incremental Cost-Effectiveness Ratio (ICER): the additional cost per beneficiary HH of	Arm 2 and Arm 3, each relative to Arm 1 (control arm) to achieve each % increase in	non-wasting prevalence (measured by WHZ). This is a ratio of incremental cost (cost per	household indicator) to incremental effectiveness (% change in non-WHZ-wasted	children).

*Societal Costs*

To evaluate the required beneficiary time investment for participating in the program, societal costs were analyzed and costed using the ‘average income per capita in Somalia’^3^:

Arms 1 and 3 Time Investment: the travel and participation time required for registration and nutrition sensitization sessions for all beneficiary HHs.

Arm 2 Time Investment: travel time and participation time of lead and non-lead mothers were estimated for all M2M groups, lead mother training, cooking demonstrations, and individual counseling sessions, as well as the cash transfer related registration requirements included for Arm 1 and 3.

**Appendix S10. Market Monitoring Data Collection Sites**

| **Region** | **Market Monitoring Data Collection Areas** |
| --- | --- |
| *Bay* | Barxada  Hanano  Ondon  Adaada  Dugandug  Baidoa |
| *Hiraan* | Yoobsan/daraawiish  Jawil  Mahas  Gerjiir  Beegadid  Mataban |

**Appendix S11. Food Commodities Monitored During Market Monitoring**

| **Category** | **Foods** |
| --- | --- |
| *Sentinel Foods* | Sorghum flour, white rice, pasta, wheat flour, imported rice, vegetable oil |
| *Fruits and Vegetables* | Banana, Swiss chard, groundnuts, pumpkin, spinach, potatoes, lime, onion |
| *Cereals* | Beans, lentils, maize, whaite maize, sorghum, red sorghum, cowpeas, baits (canned) |
| *Protein Sources* | Meat, eggs, cooked eggs, goat liver, canned sardines, canned milk, camel meat, camel milk |

**Appendix S12. Effectiveness and Cost-Effectiveness of Cash-Based Interventions on Nutrition Outcomes**

Cash-based interventions (CBIs) have been shown to be more effective at decreasing food insecurity and malnutrition, especially in low- and middle-income countries and humanitarian contexts.[1] CBIs can vary, ranging from straightforward unconditional cash transfers to more established programs like conditional transfers or voucher programs. CBIs aim to enhance access to nutritious foods and general nutrition. CBIs may also be employed to achieve broader health outcomes related to maternal and child health, such as wasting and stunting.

This literature review discusses the impacts of CBIs on humanitarian and low-income nutritional outcomes in the form of malnutrition reduction, dietary diversity, and cost-effectiveness. Therefore, a targeted review of five studies and one report was employed to examine conditional and unconditional cash transfers in environments with high food insecurity. Studies were chosen based on their nutritional effect assessments and relevance to CBIs conducted in humanitarian or low-income settings.

The studies examined programs in Pakistan, Syria, Niger, and Burkina Faso with various CBI designs. In Niger, one study compared the costs, cost-effectiveness, and cost-efficiency of two cash-based interventions for preventing acute malnutrition in children under five years. It compared a modified cash transfer program, which began earlier and ran longer to a standard program.[2] In Pakistan, a trial evaluated the impact of three CBIs (standard cash, double cash, and fresh food vouchers) on child nutrition outcomes in a humanitarian context. It considered the effects on stunting and wasting at six months and one year and on enhancing nutritional resilience.[1,3] In Burkina Faso, research was concerned with the cost and cost-effectiveness of mobile cash transfers for the MAM'Out project, adopting a societal perspective as it included costs falling on implementing partners, as well as beneficiary households.[4] A study in Northern Syria examined the feasibility of CBIs, their acceptability, infrastructure, capacity for implementation, value for money, risks, and responsiveness to the needs of beneficiaries. The study aimed to guide the way forward in humanitarian aid approaches and steer the scale-up of cash-based modality in a context historically dominated by in-kind programming.[5]

This review highlights key parameters of intervention effectiveness, including program design and education components, as well as external factors such as conflict and displacement.

**Overall Effectiveness of CBIs on Nutrition Outcomes**

Across several studies, there is a growing consensus that cash transfers can improve nutrition outcomes, particularly for children and mothers. However, their effectiveness varies based on factors such as transfer modality, conditionality, program duration, and complementary support services. Studies from Sindh Province, Pakistan, and Tapoa Province, Burkina Faso, both humanitarian settings where CBIs were used to address food insecurity. In Pakistan, a trial assessed the impact of unconditional cash transfers: Standard Cash (SC), Double Cash (DC), alongside Fresh Food Vouchers (FFV) on child nutrition.[1,3] In Burkina Faso, research focused on the cost-effectiveness of mobile unconditional cash transfers under the MAM’Out project, with community sensitization sessions encouraging spending on food.[4] By comparing these approaches, this review explores how cash transfer modalities and program structures influence nutrition outcomes in humanitarian settings.

One key finding from successful CBI programs is their impact on dietary diversity. Unconditional cash transfers often enable households to improve their food purchasing, leading to higher-quality diets. This is particularly important for households facing extreme poverty or seasonal food shortages.[4] For instance, a study conducted in Pakistan, a cluster-randomized controlled trial evaluating different CBIs, assessed the effects of SC, DC, FFV on maternal and child nutrition outcomes.[1] While it was initially hypothesized that FFVs would have the greatest impact on child growth and micronutrient status by increasing dietary diversity, the results showed a more significant improvement in dietary diversity for the DC arm, followed by SC, with FFV showing the least improvement. However, in terms of weight-for-height z-scores (WHZ), children in the FFV arm experienced greater growth compared to those in the SC arm.[1]

According to evidence from child nutrition studies, cash transfers can help reduce stunting and wasting in children under the age of five, especially when accompanied by nutrition instruction or counseling^4^. In Burkina Faso, the MAM'Out initiative found that conditional cash transfers combined with nutrition instruction had a considerably higher impact on child nutrition outcomes than programs that offered neither cash nor educational support^4^. This emphasizes the need of combining financial support with SBCC measures to increase the effectiveness of CBIs in combating malnutrition.[4]

CBI success is affected by a range of programmatic and environmental factors. Cash transfer amounts, intervention duration, and whether the program has complementary services (e.g., health access, nutrition counseling) all impact results. Modality of transfer and delivery mechanisms are also crucial. Mobile cash transfer mechanisms, for example, can be used to boost access in areas with fewer banks, while direct cash transfers may prove more effective in settings where digital payment infrastructure is low.[1,4] It is therefore important to consider these context-specific factors to optimize CBIs and ensure that they become effective in improving food security and reducing malnutrition among the target groups.

**Cost-Effectiveness of CBIs**

Understanding the cost-effectiveness of CBI’s is crucial, particularly when compared to in-kind food aid. This review examines cost-effectiveness by analyzing peer-reviewed studies from Burkina Faso, Syria, and Pakistan, along with an organizational report from the Cash Alliance on emergency cash support in Somalia. The Burkina Faso study assessed the cost-efficiency of mobile unconditional cash transfers, while research in Syria examined the feasibility and value for money of CBIs in a conflict-affected region.[4,5] A Pakistan study evaluated the cost-effectiveness of SC, DC, and FFV in improving child nutrition outcomes.[3] Findings suggest that while CBIs can be a cost-effective alternative to in-kind aid, efficiency varies based on delivery mechanisms and impact per dollar spent.

CBIs are often more cost-efficient than in-kind food assistance due to lower logistical and operational costs.[5] Unlike food aid, cash does not require extensive infrastructure for storage, transportation, or distribution, making it a more scalable option, particularly in humanitarian settings where resources are limited.[5] Mobile transfers further enhance efficiency by reducing costs associated with manual cash distribution while ensuring direct payments to recipients, minimizing intermediary handling and corruption risks.[5] For instance, mobile cash transfers in Somalia allowed beneficiaries to receive funds even if they relocated, making them particularly beneficial for internally displaced persons.[6] While cash transfers were generally more cost-effective, the specific delivery mechanism had an impact on efficiency, with mobile transfers proving to be a cost-saving option to traditional distribution techniques. However, one study also revealed that cost-efficiency is not universal across all CBIs. A study in Dadu, Pakistan, which evaluated the cost-efficiency of three CBI models: SC, DC, and FFV, found that cash beneficiaries incurred significantly higher personal costs compared to voucher recipients.[3] Unlike voucher distribution, which took place directly in beneficiaries’ villages, most cash recipients had to travel to collect their payments, sometimes spending an entire day due to public transportation schedules.[3] This suggests that while CBIs can reduce overall program costs, their design must consider beneficiary accessibility to avoid unintended financial burdens.

Several studies have found that CBIs, particularly unconditional cash transfers, provide better returns on investment in terms of nutrition intake and improvement.[1,3,4] The flexibility of cash allows recipients to allocate funds according to their specific needs, maximizing the effectiveness of each dollar spent. In contrast, food aid and voucher systems impose spending restrictions, which can limit their overall cost-effectiveness by preventing households from prioritizing essential nutritional needs.[5]

However, financial flexibility alone does not guarantee improved dietary outcomes. While CBIs allow households to allocate resources according to their needs, their impact depends on market access and recipient knowledge. If beneficiaries lack nutrition education or access to affordable and diverse food options, the benefits of cash transfers may be limited. Globally, the way markets function in crises remains poorly understood, and the comparative impact of different humanitarian assistance modalities on market dynamics is still an area of ongoing research.[5] Market disruptions, inflation, or supply shortages can reduce the purchasing power of cash assistance, making it less effective than in-kind food aid in certain contexts.[5] Therefore, conducting market assessments in target areas is essential to determine the feasibility of CBIs as a substitute for traditional food assistance.

**Specific Interventions and their Impacts**

The studies show that the design of cash interventions has a major impact on their effectiveness. Some of the intervention models include:

*Unconditional Cash transfers (UCTs)*

UCTs provide recipients with financial support without strings attached, and the freedom to use it as per immediate needs. They are greatly appreciated in humanitarian settings for the timeliness of their rollout and for the dignity of allowing recipients to make their own financial decisions. However, while UCTs have been shown to improve food security, they may not always lead to significant health and nutrition change in children in the absence of complementary support.[3] Some limiting factors include beneficiaries' low nutritional knowledge and households' limited market access to a diversity of affordable foods, which can prevent households from making the most nutritionally effective purchases. A study in Somalia found that the combination of nutrition counseling with UCTs was more effective in improving the growth of children and household food security than UCTs alone.[7] This emphasizes the requirement to combine UCTs with complementary interventions, such as nutrition education and health.

*Conditional Cash Transfers (CCTs)*

CCTs only pay when the beneficiary meets specified conditions, such as going to health centers, participating in nutrition education classes, or enrolling in community-based behavior programs. By linking cash assistance to health and education activities, CCTs can lead to significant longer-term changes in child health, care consumption, and well-being compared to unconditional transfers.[5] These programs promote health-seeking behaviors, and the beneficiaries are able to receive the required services that can enhance their nutritional status.[5] While CCTs have demonstrated stronger impacts on long-term health and education, their success depends on program design, implementation, and access to required services. In settings where health facilities or nutrition programs are not readily available, it may be hard to achieve CCT requirements, limiting the reach and impact of these interventions.[5]

*Double Cash (DC)*

DC transfers involve providing double the value of cash relative to standard cash transfers, which can be conditional or unconditional depending on program design. While not necessarily targeted for emergencies, DC transfers can be particularly valuable during peak-demand times of food insecurity, such as seasonal hunger gaps or crises, by providing households with increased financial flexibility to meet short-term food and nutritional needs.[2,5] None of the studies reviewed specifically assessed resilience indicators, such as the Reduced Coping Strategies Index (rCSI) or household debt levels, in relation to DC transfers.

*Cash + Complimentary Services*

Cash + Complimentary services also known as "Cash Plus" models combine cash transfers which can be conditional or unconditional with additional support services such as nutrition education, social and behavior change communication, healthcare, or agricultural assistance. Unlike CCTs, which require recipients to meet specific conditions to receive cash, Cash Plus programs provide integrated support to address barriers beyond financial constraints. By addressing multiple determinants of nutrition, these multifaceted initiatives frequently have the greatest impact on nutrition outcomes, particularly in areas where knowledge about dietary choices is limited or access to nutritious food is restricted.[4]

**Educational Components of Cash-Based Interventions**

Education interventions and sensitization campaigns play a central role in complementing the impact of CBIs on the nutrition of children. Behavior Change Communication (BCC) training and community sensitization are the most common approaches, both aimed at improving caregiver choice-making and education regarding nutrition, hygiene, and child growth.[2] The education component is usually bundled with CBI interventions to confirm healthy health habits and optimize the impact of the cash transfer.

BCC Sessions are usually conducted in conjunction with cash transfers or other food interventions to provide caregivers with functional nutrition and health education, covering topics such as optimal breastfeeding, complementary feeding practices, hand washing, and bed net use. Cooking demonstrations are sometimes conducted on-site to enable caregivers to prepare healthy meals from resources available to them.[1]

As a broader form of Behavior Change Communication (BCC), community sensitization supports CBIs by raising awareness among beneficiaries about the importance of allocating cash toward child nutrition and development. These sessions fill the gap between cash provision and behavior change towards better nutrition outcomes.[4]

A good example of an intervention that integrated BCC and community sensitization was the WINS Program in Sindh Province, Pakistan, in a humanitarian setting. All study participants received monthly group education by research mobilizers.[1,3] Education covered important topics such as:

- Causes of undernutrition
- Benefits of exclusive breastfeeding
- Improved complementary feeding practices
- Food and water hygiene
- Handwashing and sanitation

In addition to micronutrient supplementation and outpatient treatment of severely malnourished children, the WINS program also aimed to educate caregivers with information required to maintain child nutrition and health.[1] With the integration of BCC and community sensitization into CBIs, these interventions can enhance the long-term impact of cash transfers on child development and health.

**Gaps Identified in the Literature on Cash-Based Interventions for Child Nutrition**

Though cash plus interventions have improved nutritional outcomes compared to cash alone, few of the studies fail to quantify major determinants that lead to malnutrition. Some studies quantify child nutrition status using anthropometric measures of weight-for-height z-scores (WHZ) and height-for-age z-scores (HAZ), but neither household-level food security nor dietary diversity is quantified.[1,3] None of the studies made use of Food Consumption Scores (FCS) and Household Dietary Diversity Scores (HDDS), constraining our ability to identify how household food consumption habits and food availability are affected by economic interventions. Consistency in the measurement of the anthropometric indicator was also absent between the studies, making comparative analysis challenging. Closing these gaps by introducing household-level food security and dietary diversity indicators in addition to the standard anthropometric ones would contribute more to greater comprehension of CBIs' child nutrition gains over immediate growth-related ones.

Nearly all studies evaluate the short- to medium-term impacts of cash-based interventions on child nutrition and household outcomes and were typically less than one year in length. Some nutrition markers, such as wasting, may react relatively swiftly, while others, such as stunting, require longer intervention before substantial change is noted. There is little data, though, on long-term sustainability of gains following interventions. Whether decreases in stunting and wasting or in dietary variety are maintained long term after the discontinuation of cash transfers is unknown. Further studies are needed to assess the long-term effects of CBIs on child nutrition.

**Emerging Research and Protocols in Cash-Based Interventions**

There are many emerging studies being proposed and conducted pertaining to cash-based interventions. Recent studies have also published research protocols to evaluate cash-based interventions (CBIs). [8,9] Although most are still in early stages without final outcomes, their designs highlight essential factors for child and maternal nutrition, such as differences in cash delivery methods, program duration, and integration of additional support services. Each study has a unique approach in investigating the impact and challenges of CBIs:

- Huda (2020)[8]: The study, implemented in rural Bangladesh, focuses on conditional cash transfers connected to maternal healthcare visits, evaluating the impact of certain health behaviors, such as exclusive breastfeeding, maternal nutrient intake, and complementary feeding practices can boost nutritional outcomes. It measures dietary improvements and long-term health indicators like stunting and wasting, exploring how health education might directly influence behavior changes ^8^
- Martin-Canavate (2023)[9]: This protocol paper aims to assess the cost-effectiveness and cost-efficiency of three interventions to reduce stunting in children under 2 from low-income households in Cunene and Huila, Angola. The interventions include: (1) standard of care (SoC) alone, (2) SoC plus nutritional supplements, and (3) SoC plus cash transfers. The economic evaluation will take a societal perspective to estimate implementation costs for each intervention and costs for participants and families. It will calculate cost-efficiency per woman, child, and household reached and estimate cost-effectiveness by determining the cost per case of stunting averted compared to standard care.

| **Appendix S13. Strengths and Limitations of Cost Analysis Approach** | |
| --- | --- |
| Strengths | In-depth understanding: The KIIs and focus group discussions allowed for in-depth exploration of the newly introduced concept of societal opportunity costs, which is often difficult to replicate capture through independent quantitative surveys. Furthermore, programme staff often possess nuanced understanding of the context and can provide rich qualitative data on how the intervention affects beneficiaries' choices and behaviours. |
|  | Contextualized information: The discussions also provided valuable contextual information about the specific circumstances in Hiran and Bay, such as economic realities, and social dynamics, which influence opportunity costs. |
|  | Access to insider perspectives: SCI staff have direct experience of the intervention and its impact on beneficiaries, and they were able to offer insights into the unintended consequences and hidden costs associated with participation in the programme, where relevant, which may not have been readily apparent. |
|  | Flexibility and adaptability: The KIIs and focus groups also allowed for flexibility in probing emerging themes and exploring unexpected responses, leading to a richer understanding of the factors affecting opportunity costs and the wider economic evaluation. |
|  | Triangulation: Information gathered from the discussions were then used to triangulate and validate data collected through other methods (e.g. direct economic costings), thereby increasing the reliability of the wider cost-efficiency findings. |
| Limitations | Subjectivity and bias: As with any qualitative method, participants (in this case, SCI staff) may have inherent biases due to their involvement in the intervention. As such, they may downplay negative consequences, such as societal impacts, potentially skewing the estimation of opportunity costs. |
|  | Limited generalizability: Findings from KIIs and the focus groups may not be representative of the wider population in Hiran and Bay. The perspectives of programme staff may not reflect the diverse experiences and viewpoints of all beneficiaries. |
|  | Difficulty in quantification: While interviews and focus groups can provide valuable qualitative insights, it can be challenging to translate these qualitative narratives into quantifiable measures of opportunity costs. This limitation can hinder precise estimation and comparison of costs across the different R2HC Arms and regions. |
|  | Time and resource intensive: Conducting in-depth interviews with multiple programme staff is resource-intensive and highly time sensitive to coincide with the convening of the Nairobi event, as such, there was limited sample size of participants, potentially affecting the generalizability of findings. |
|  | Availability of secondary data: Utilizing existing secondary data sources was required for estimating the economic cost of beneficiaries’ time, which could have been used for other activities (such as paid work). Sources like labour market surveys, economic reports from governmental or non-governmental organizations, and academic studies to provide data on a realistic hourly wage for the rural Somalia informal market were extremely limited. As such, a combination of the ‘Somalia average income per capita (annual)’ from a World Data source[1] and Clockify’s ‘average number of working hours per year (Somalia)’[2] was used to estimate an hourly cost. Using this highly approximate method to quantify the opportunity costs of time spent participating in the intervention will have had a limiting factor on the accuracy and reliability of the societal cost estimations. |
| Mitigation Approach | Participant selection: Due to the convening of the Nairobi event, the SCI programme informants available were ideally placed to have diverse roles and experiences within the programme to capture a range of perspectives. |
|  | Use of structured guides: To develop a standardized approach to measuring societal costs, guides were developed with clear and specific questions to ensure consistency and reduce bias (attached in email). |
|  | Triangulation of data sources: The combination of the KIIs and focus group discussions, with secondary data from programme documentation to validate findings and enhance the reliability of estimations. For example, the SCI programme staff were able to confirm the hourly wage estimate (sourced from secondary data) was realistic for the intervention context. |
|  | Quality facilitation: All KIIs and focus groups were facilitated by the lead economic evaluator. |
